# CohortContrast: An R Package for Enrichment-Based Identification of Clinically Relevant Concepts in OMOP CDM Data

**DOI:** 10.64898/2026.04.22.26351461

**Authors:** Markus Haug, Neeme Ilves, Nikita Umov, Hendrik Šuvalov, Kerli Mooses, Sirli Tamm, Helene Loorents, Marek Oja, Sulev Reisberg, Jaak Vilo, Raivo Kolde

**Author notes:** **Corresponding author:** Markus Haug, Postal address: Arvutiteaduse instituut, Tartu Ülikool, Narva mnt 18, 51009 Tartu, Estonia.

## Abstract

**Objective:** To address the unresolved bottleneck of selecting cohort-relevant clinical concepts for treatment trajectory analysis in observational health data, we introduce CohortContrast, an OMOP-compatible R package for enrichment-based concept identification, temporal and semantic noise reduction, and concept aggregation, enabling cohort-level characterization and downstream trajectory analysis.

**Materials and Methods:** We developed CohortContrast and applied it to OMOP-mapped observational data from the Estonian nationwide OPTIMA database, which includes all cases of lung, breast, and prostate cancer, focusing here on lung and prostate cancer cohorts. The workflow combines target-control statistical enrichment, temporal/global noise filtering, hierarchical concept aggregation and correlation-based merging, with optional patient clustering for downstream trajectory exploration. We validated the approach with a clinician-based plausibility assessment of extracted diagnosis-concept pairs and evaluated a large language model (LLM) as an auxiliary filtering step.

**Results:** We analyzed 7,579 lung cancer and 11,547 prostate cancer patients. The workflow reduced concept dimensionality from 5,793 to 296 concepts (94.9%) in lung cancer and from 5,759 to 170 concepts (97.0%) in prostate cancer, and identified three exploratory patient subgroups in both cohorts. In a plausibility assessment of 466 diagnosis-concept pairs, validators rated 31.3% as directly linked and 57.5% as indirectly linked.

**Discussion:** CohortContrast reduces manual concept curation by prioritizing and aggregating cohort-relevant concepts while preserving clinically interpretable treatment patterns in OMOP-based real-world data.

**Conclusion:** CohortContrast enables scalable reduction of broad OMOP concept spaces into clinically interpretable, cohort-specific representations for exploratory trajectory analysis and real-world evidence research.

## BACKGROUND AND SIGNIFICANCE

Real-world data (RWD) has become increasingly important for longitudinal analyses of patient treatment trajectories and adherence to clinical protocols, supporting evaluation of therapeutic interventions.[1] Recent work demonstrates its value across domains including breast cancer,[2] chronic obstructive pulmonary disease and medication optimization.[3,4]

However, a persistent bottleneck in such analyses lies in accurately identifying cohort-relevant clinical concepts in high-dimensional data and aggregating them into interpretable states (i.e., concept sets used as trajectory nodes). Aggregating related concepts and filtering spurious or non-specific signals are essential for meaningful phenotyping, but this often requires substantial clinical expertise and familiarity with local coding practices.[5, 6] A practical strategy is to combine top-down clinical knowledge with bottom-up data-driven prioritization.[7] Many studies still rely on custom code developed from scratch, leading to variable configurations, local dependencies and reduced reproducibility and scalability.[8] Common data models (CDMs) help address these limitations by harmonizing data structures and enabling standardized analytical tools and federated research.[6]

Our prior work has focused on developing tools for constructing treatment trajectories within the Observational Medical Outcomes Partnership (OMOP) CDM framework and facilitating federated research to investigate these pathways.[9–11] The OMOP CDM supports transparency, reproducibility and transferability through standardized data structures and analytical tooling.[12–14] Although previously published packages are effective for trajectory construction, they rely on hand-picked cohort-relevant concepts that are needed to derive clinically meaningful states for phenotyping. Given the breadth of research questions on patient trajectories in CDMs, more automated exploratory tools are needed to identify phenotype-related concepts within specific cohorts and support targeted investigations.[15, 16]

In this context, we present CohortContrast, an OMOP-compatible framework for exploratory identification of cohort-relevant clinical concepts through statistical enrichment analysis. The approach retrospectively contrasts target and control cohorts using relative risk and statistical significance metrics. Conceptually, it is similar to enrichment-based knowledge discovery in genomics, where enrichment approaches prioritize relevant signals from high-dimensional inputs.[17, 18] To further reduce the number of concepts the method performs semantic aggregation and abstraction by leveraging ontology structure (e.g., SNOMED CT for condition concepts).[19] We demonstrate CohortContrast on two large oncology cohorts, showing substantial concept-space reduction while preserving clinically interpretable signals to support downstream trajectory analyses.

## METHODS

Within the OMOP community, large-scale observational analyses often use reusable R workflows over OMOP-mapped data, with standardized cohort definitions commonly built in ATLAS to support transparency, reproducibility and transferability.[20] We introduce CohortContrast, an R package that adds an exploratory cohort-characterization step to the OMOP CDM workflow. By incorporating data-driven concept discovery, CohortContrast identifies cohort-relevant clinical concepts without relying exclusively on pre-specified phenotype definitions.

All methods described in this study are implemented in the CohortContrast package, which is available from the Comprehensive R Archive Network (CRAN) and GitHub.[21, 22] The principal input is a target cohort specifying patients and observation windows, contrasted against user-defined control periods or cohorts. CohortContrast also includes a graphical user interface (GUI) for configuration, result exploration and optional clustering. For sharing results, a small-cell suppression module is implemented.

The CohortContrast workflow (Figure 1; Appendix 1) consists of four phases: cohort definition (Phase I), statistical enrichment (Phase II), temporal-bias filtering using a matched cohort comparison (Phase III) and hierarchy- and correlation-based concept aggregation (Phase IV).

**Figure 1.**
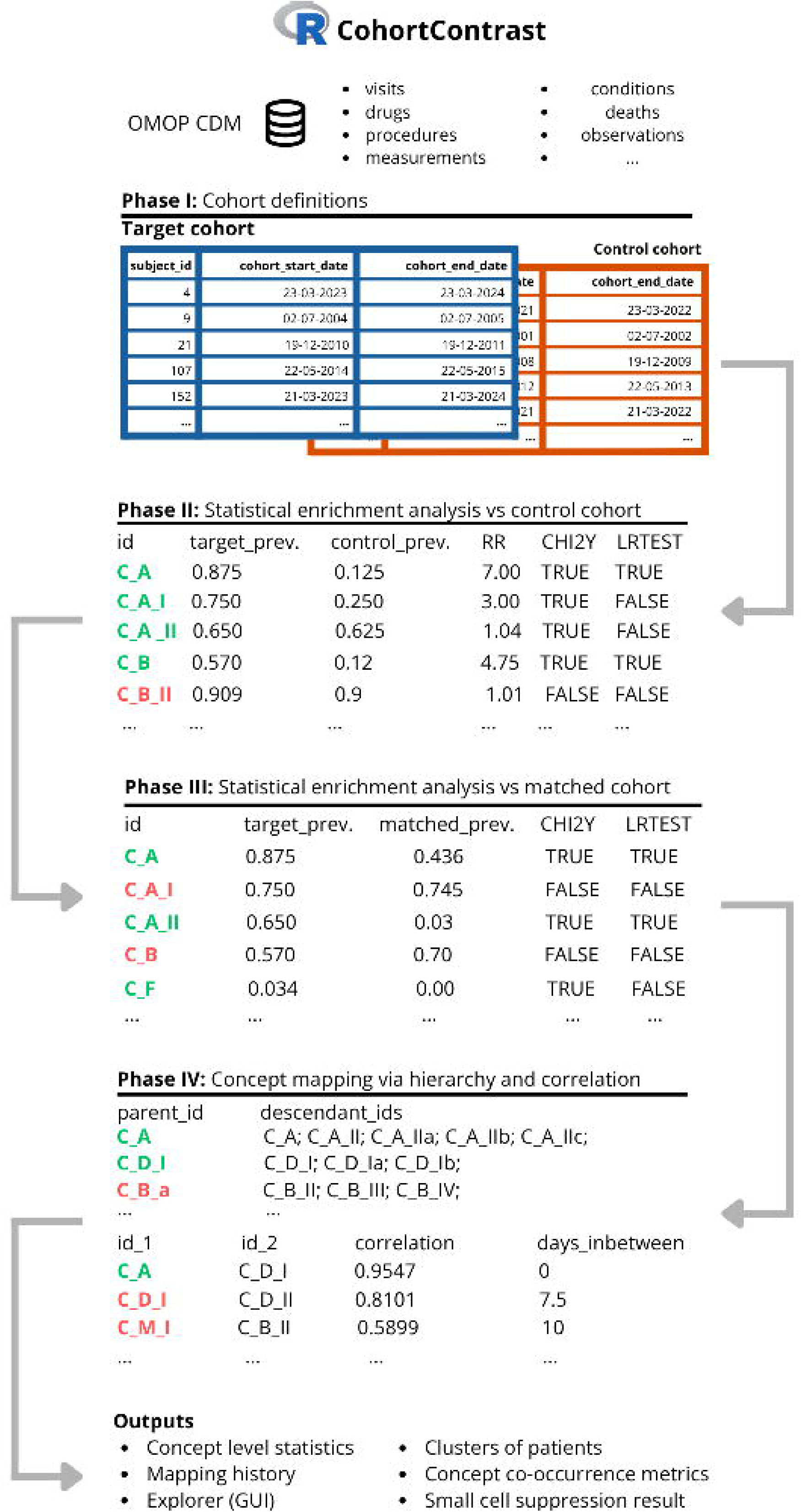
The workflow of the CohortContrast package comprises four phases. The first phase is for target and control cohort definition. The second phase filters for concept enrichment illustrated by risk ratio (RR) using CHI2Y and LRTEST. The third phase removes temporal bias using CHI2Y and LRTEST. The fourth phase maps descendant concepts together with present parents also combining frequently co-occurring concepts via correlation.

### Phase I: Cohort definitions

The primary input to CohortContrast is a target cohort, defined by the patients of interest together with the corresponding observation windows (cohort start and end dates). These windows can be specified to represent different clinically meaningful periods, such as post-diagnosis treatment, post-intervention follow-up, or prediagnostic intervals. Clinical concepts (standard concepts over all domains) recorded within the target observation windows are treated as candidate concepts for downstream enrichment analysis.

To enable cohort contrast, the package supports multiple control strategies, including self-controlled comparisons (using alternative time windows from the same patients) and matched control cohorts (eg, age- and sex-matched individuals). The target and control definitions established in this phase provide the basis for statistical enrichment testing in Phase II, where concepts overrepresented in the target cohort are identified.

### Phase II: Statistical enrichment analysis

The OMOP CDM contains a large number of standardized clinical concepts,[23] making it challenging to identify which concepts are most relevant for a given cohort. In this phase, CohortContrast identifies concepts that are statistically overrepresented in the target cohort relative to the control cohort using two complementary tests: a chi-squared test for two proportions with Yates continuity correction (CHI2Y) and univariate logistic regression (LRTEST; Wald test). CHI2Y can be conservative under sparse counts, whereas LRTEST can be more sensitive but is non-estimable under (quasi-)complete separation. We used an OR rule across CHI2Y and LRTEST because the two tests are complementary with respect to their failure modes. Similar approaches have been used in feature selection for predicting COVID-19 mortality and emergency readmissions.[24, 25] These methods are computationally efficient and interpretable, making them suitable for large-scale observational datasets.

For each clinical concept, a binary patient-level indicator is constructed to represent whether the concept is observed at least once within the defined observation window. Concept prevalence is compared between target and control cohorts using CHI2Y and LRTEST. Risk ratios (RR) are also computed as effect-size summaries of enrichment. Concepts are retained as enriched if they are more prevalent in the target cohort and meet a Bonferroni-adjusted significance threshold of 0.05 (over all concepts) in either CHI2Y or LRTEST. This dual-test filtering strategy reduces retention of broadly prevalent or non-discriminative concepts while preserving cohort-specific signals for downstream analysis.

### Phase III: Temporal bias mitigation

Because observation windows are user-defined, enrichment results may be influenced by calendar-time effects and other temporal background signals unrelated to the target cohort. To address this, CohortContrast provides an optional third phase that repeats the enrichment analysis against a covariate- and time-matched global cohort drawn from the same data source. By default, we perform 1:1 matching on sex and age, and we align calendar time by assigning each matched comparator the same index date and exit date as its paired target patient; the matching ratio and covariates are configurable.

For each concept, we apply CHI2Y and LRTEST to compare concept prevalence between the target cohort and the matched global cohort. Concepts statistically not more prevalent in target are removed from the Phase II output to improve specificity of the retained concept set. For example, when observation windows overlap the years 2020–2022, COVID-19-related concepts may appear enriched in many cohorts within comprehensive healthcare datasets due to global temporal effects rather than disease-specific treatment patterns. Filtering such concepts reduces the influence of non-specific temporal background signals on downstream interpretation.

### Phase IV: Hierarchy- and correlation-based concept aggregation

After statistical enrichment, the retained output often includes overlapping or semantically similar concepts, which can hinder interpretation and downstream analysis. To improve clarity and reduce redundancy, CohortContrast applies a two-step aggregation procedure that groups closely related clinical concepts while preserving their clinical meaning.

First, the method leverages hierarchical relationships in standardized OMOP vocabularies to perform hierarchy-based aggregation. When a retained parent concept and one or more retained descendant concepts are both present, descendant concepts are merged into the parent to reduce unnecessary granularity. To preserve interpretability, we retain a child concept rather than merging it when the child’s prevalence exceeds that of its parent. When multiple nearest retained ancestors are available, we select the most prevalent ancestor as the aggregation target. This procedure is applied across domains by merging each retained concept to its nearest retained ancestor in the vocabulary hierarchy. Similar hierarchical aggregation strategies have been described by Willett et al.[26] and are widely used in the OHDSI community.[20]

Second, we perform greedy correlation-based aggregation within each OMOP domain to merge concepts with similar occurrence patterns. Following prior work on redundant SNOMED CT concepts,[27] we compute Pearson correlations from binary patient–concept presence matrices and flag pairs with strong co-occurrence (r > 0.7). We then require a median absolute temporal offset of 0 days: for each patient with both concepts present, we compute all pairwise absolute differences (in days) between occurrence dates of the two concepts and take the median as the patient-level offset (set to infinity if dated occurrences are missing for either concept); the concept-pair offset is the median of these patient-level offsets across patients. Pairs meeting both criteria are merged, and merged concepts are labeled by joining names with “+”, reducing redundancy for downstream cohort characterization and trajectory analysis.

### Clustering

CohortContrast includes an optional, exploratory post-processing step to cluster patients based on enriched concepts and their temporal patterns (Appendix 2). We construct a patient-by-concept feature matrix from retained concepts using occurrence indicators and timing relative to the cohort index date. Features are standardized and reduced using principal component analysis (PCA), and patients are clustered using k-medoids in the PCA space. Candidate values of k (default 2–5) are evaluated using silhouette scores.[28] Similar RWD clustering approaches have been reported previously.[29–31]

### Visualization

To facilitate interpretation of the reduced longitudinal concept space, we developed a composite visualization that integrates temporal, epidemiologic, demographic, and cluster-level summaries within a single figure (Figures 3–4). Concepts are faceted by hierarchical lineage and ordered by the median timing of first occurrence relative to the index date, enabling temporal structure to be assessed alongside semantic grouping. For each concept, the display includes (i) violin and box plots summarizing the distribution of event timing across the observation window, (ii) bar plots showing cohort prevalence and enrichment relative to controls, interval plots quantifying deviations in (iii) age and (iv) sex from cohort averages, and (v) a heatmap of cluster-specific prevalence and median occurrence counts. Together, these components provide a compact and interpretable overview of the concepts that characterize the target cohort.

### Case study

To evaluate the workflow, we applied CohortContrast to lung and prostate cancer cohorts constructed in an OMOP CDM instance derived from the Estonian nationwide OPTIMA Consortium database, transformed to OMOP CDM following the protocol and data quality procedures described by Oja et al.[32]. OPTIMA integrates claims (provided services and costs), prescription data, electronic health records, mortality data, and the Estonian Cancer Registry for patients diagnosed with lung, breast, or prostate cancer in Estonia between 2012 and 2022. Across the database, 63,757 individuals contribute 12,767 unique standardized clinical concepts.

For each cancer type, cohorts were defined using the prevalent diagnosis concepts malignant tumor of lung (SNOMED CT: 363358000) and malignant tumor of prostate (SNOMED CT: 399068003), including descendant concepts defined through the OMOP vocabulary hierarchy. Inclusion required at least a three-year lookback period with no prior documentation of the corresponding cancer in condition occurrences to identify first-time cases (Figure 2).

**Figure 2.**
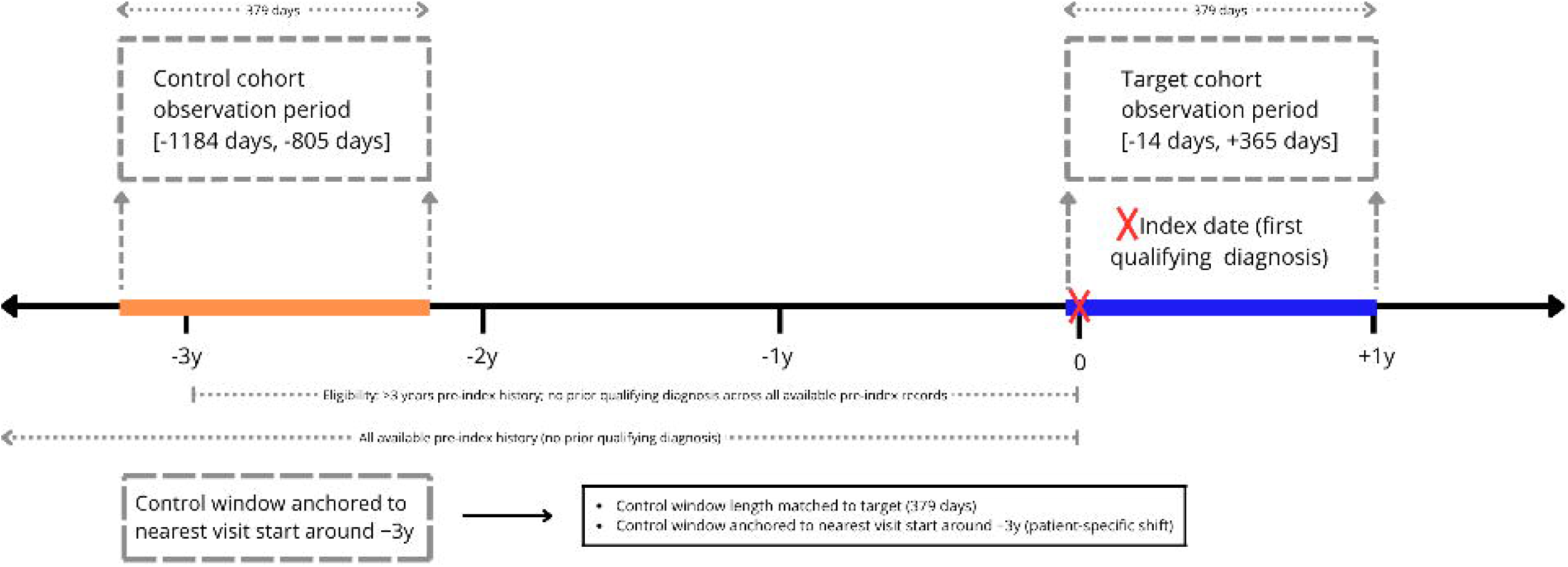
Target and control cohorts’ observation periods to the baseline anchor date (i.e., three years prior to diagnosis).[33] The resulting lung cancer cohort included 7,579 patients and the prostate cancer cohort included 11,547 patients (Appendix 3).

Patients were followed for one year post-diagnosis, with an additional 2-week pre-index interval to capture early diagnostic concepts. We used a self-controlled design in which each patient served as their own control by contributing data from the period three to two years prior to diagnosis as the baseline for enrichment comparison. To reduce bias from differential encounter density, we aligned each patient’s control window to begin at the start of the clinical visit closest

### Validation

To assess the clinical plausibility of automatically derived diagnosis–concept relationships, we conducted an independent multi-rater evaluation on all extracted diagnosis–concept pairs. Two medical doctors (MD1 and MD2) independently labeled each pair as *direct*, *indirect*, or *noise* using a predefined rubric. In parallel, we evaluated the feasibility of partially automating this step by prompting the Gemini model (gemini-3-pro-preview; temperature = 0; zero-shot) to assign the same labels and collecting structured outputs for analysis.[34]

All raters were provided identical inputs and instructions (Appendix 4), clinician raters were blinded to one another’s assessments. Concordance was quantified using pairwise percent agreement and unweighted Cohen’s _κ_ across the three categories for MD1–MD2, MD1–LLM, and MD2–LLM comparisons.[35] As an auxiliary large-scale screen, we additionally applied the same LLM labeling procedure to concepts excluded during the workflow to identify potentially relevant concepts that may have been removed during filtering.

## RESULTS

We first quantified the number of unique clinical concepts retained at each workflow phase for each cohort. After applying our workflow, we observed a substantial reduction in concept dimensionality: 94.9% for the lung cancer cohort and 97.0% for the prostate cancer cohort (Table 1). In both cohorts, the greatest reduction was observed in the condition domain, while the smallest reduction occurred in the ‘other’ domain, including death, visit and visit detail domains.

**Table 1.** Concept counts and reduction rates for lung and prostate cancer cohorts.

| Domain | Phase I | Phase II | Phase III | Phase IV | Reduction |
| --- | --- | --- | --- | --- | --- |
| Lung cancer cohort |  |  |  |  |  |
| Condition | 3087 | 150 | 89 | 44 | 98.6% |
| Procedure | 1067 | 164 | 113 | 60 | 94.4% |
| Drug | 940 | 96 | 70 | 70 | 92.6% |
| Measurement | 280 | 132 | 117 | 78 | 72.1% |
| Observation | 410 | 63 | 39 | 37 | 91.0% |
| Other | 9 | 8 | 8 | 7 | 22.2% |
| **Total** | **5793** | **613** | **436** | **296** | **94.9%** |
| Prostate cancer cohort |  |  |  |  |  |
| Condition | 3016 | 100 | 28 | 14 | 99.5% |
| Procedure | 1121 | 151 | 76 | 39 | 96.5% |
| Drug | 926 | 91 | 30 | 30 | 96.8% |
| Measurement | 274 | 114 | 67 | 52 | 81.0% |
| Observation | 414 | 66 | 31 | 29 | 93.0% |
| Other | 8 | 8 | 7 | 6 | 25.0% |
| **Total** | **5759** | **530** | **239** | **170** | **97.0%** |
| Weighted concept reduction totals per phase |  |  |  |  |  |
| **Total** | **NA** | **90.1%** | **40.9%** | **31.0%** | **96.0%** |

In Phase II, concept filtering using enrichment analysis reduced the concept space by 90.1%. Phase III achieved an additional weighted (by concept count) 40.9% reduction. Phase IV, which includes hierarchical aggregation and correlation-based mapping, further reduced the remaining concepts by a weighted average 31.0%.

As a result of the workflow for concepts across the domains, we identified a total of 296 concepts for the lung cancer cohort and 170 for the prostate cancer cohort. Clustering the patients with these concepts yielded the best silhouette scores (0.143 and 0.102) with three clusters (C1, C2, C3) for both cases.

With respect to the lung cancer clusters, we observed broadly similar demographics (C1: n=2949, mean age 69.2, 72% male; C2: n=2596, mean age 71.5, 68.7% male; C3: n=2034, mean age 68.4, 65.3% male). Despite minimal demographic separation, clusters differed markedly by recorded stage at presentation (Figure 3). C1 and C2 were enriched for advanced/metastatic staging (AJCC/UICC cM1/Stage 4: 43.0% and 34.3%), whereas C3 showed a predominantly non-metastatic/early-stage signature (AJCC/UICC cM0: 63.6%; Stage 1A: 23.7%; pN0: 36.5%), with comparatively low cM1/Stage 4 prevalence (8.3%).

**Figure 3.**
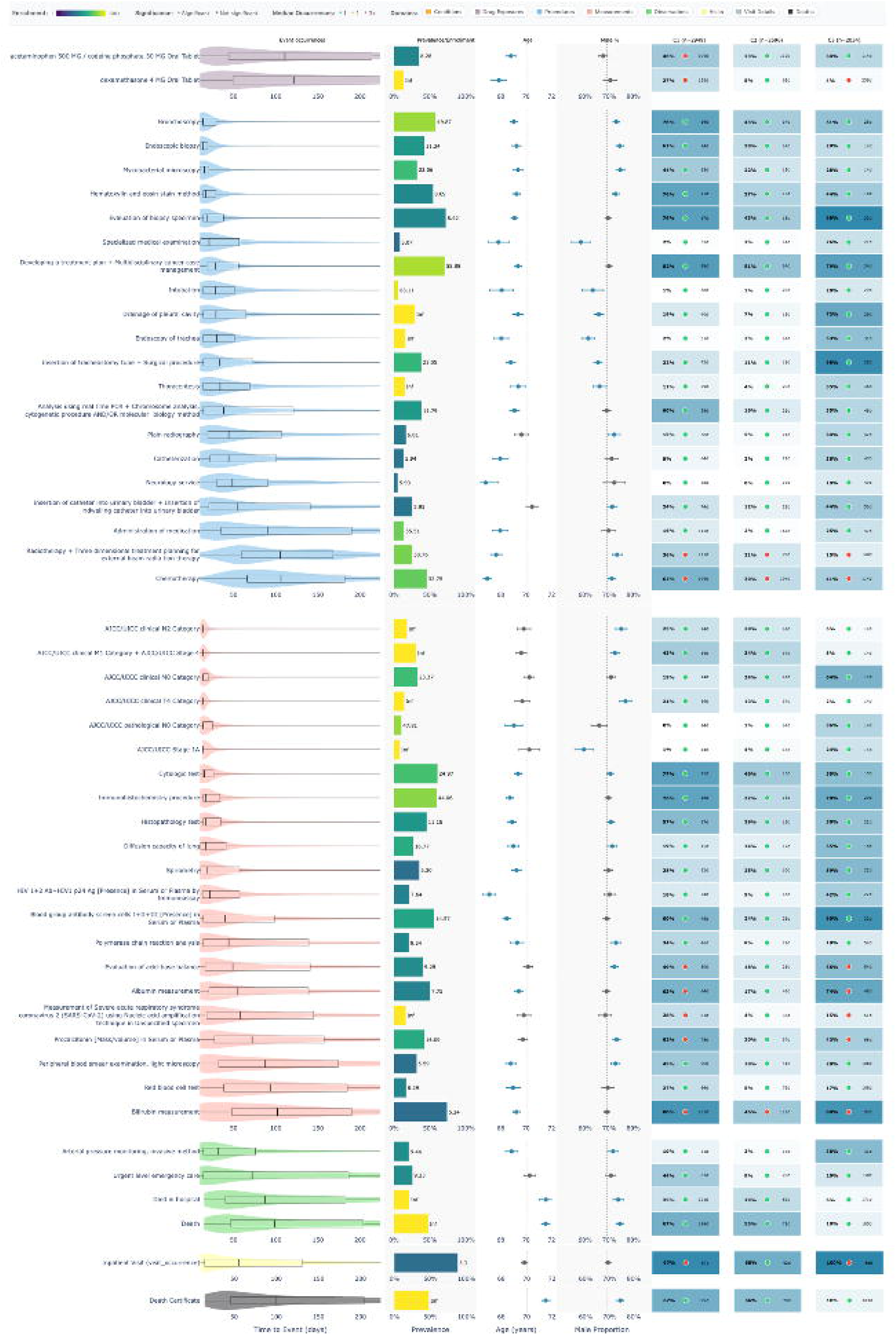
Clustering for lung cancer patients. The heatmap shows the 50 concepts with the highest between-cluster variance. Event-occurrence plots show concept timing across the observation period. The prevalence/enrichment panel summarizes prevalence in the target cohort and enrichment versus controls (Inf indicates no control occurrences). The age/male panel shows distributions and confidence intervals relative to the average. The cluster panel shows cluster-specific prevalence and median occurrence counts.

Care pathways and short-term outcomes further separated the phenotypes (Figure 3). C1 reflected the most treatment-intensive oncologic course, with higher utilization of chemotherapy (62.4%) and radiotherapy planning (36.1%), alongside supportive/adjunctive therapies (e.g., filgrastim 26.0%, dexamethasone 27.3%) and higher acute-care/infection-associated signals (urgent emergency care 44.2%, procalcitonin 61.9%). C2 exhibited a limited treatment/rapid decline pattern, with lower recorded systemic therapy (chemotherapy 30.1%) and supportive agents (filgrastim 6.4%, dexamethasone 5.4%) and the earliest mortality signal (death 54.9%), including frequent in-hospital death (29.3%). In contrast, C3 was characterized by an early, procedure and ICU heavy respiratory/pleural care signature (e.g., tracheostomy 94.8%, pleural drainage 72.4%, invasive arterial pressure monitoring 57.5%) with higher pulmonary function testing (spirometry 58.6%) and substantially lower mortality over follow-up. (Appendix 5)

For the prostate cancer cohort, demographic separation was driven primarily by age: C1 (n = 3,782; mean 70.98), C2 (n = 5,137; mean 71.25) and C3 (n = 2,628; mean 64.7). All clusters shared an early diagnostic/staging backbone around diagnosis, including biopsy of prostate and core needle biopsy using ultrasound guidance (C1/C2/C3: 48.7%/67.0%/82.0%), evaluation of biopsy specimen and H&E stain method (81.8%/71.1%/100.0%), histopathology test (21.7%/16.7%/30.4%) and immunohistochemistry procedure (39.4%/35.2%/52.7%). Clusters nevertheless differed markedly by recorded stage signature: C3 showed a strongly non-metastatic/localized pattern with high AJCC/UICC cM0 (96.3%), cN0 (62.8%), stage 2 (60.2%) and pathological T2c (38.0%), whereas C1 had the highest AJCC/UICC cM1 and stage 4 prevalence (18.4%, vs 8.3% in C2 and 1.9% in C3). C2 was intermediate by stage, with relatively higher cM0 (44.1%) than C1 (32.3%) but far below C3 (Figure 4).

**Figure 4.**
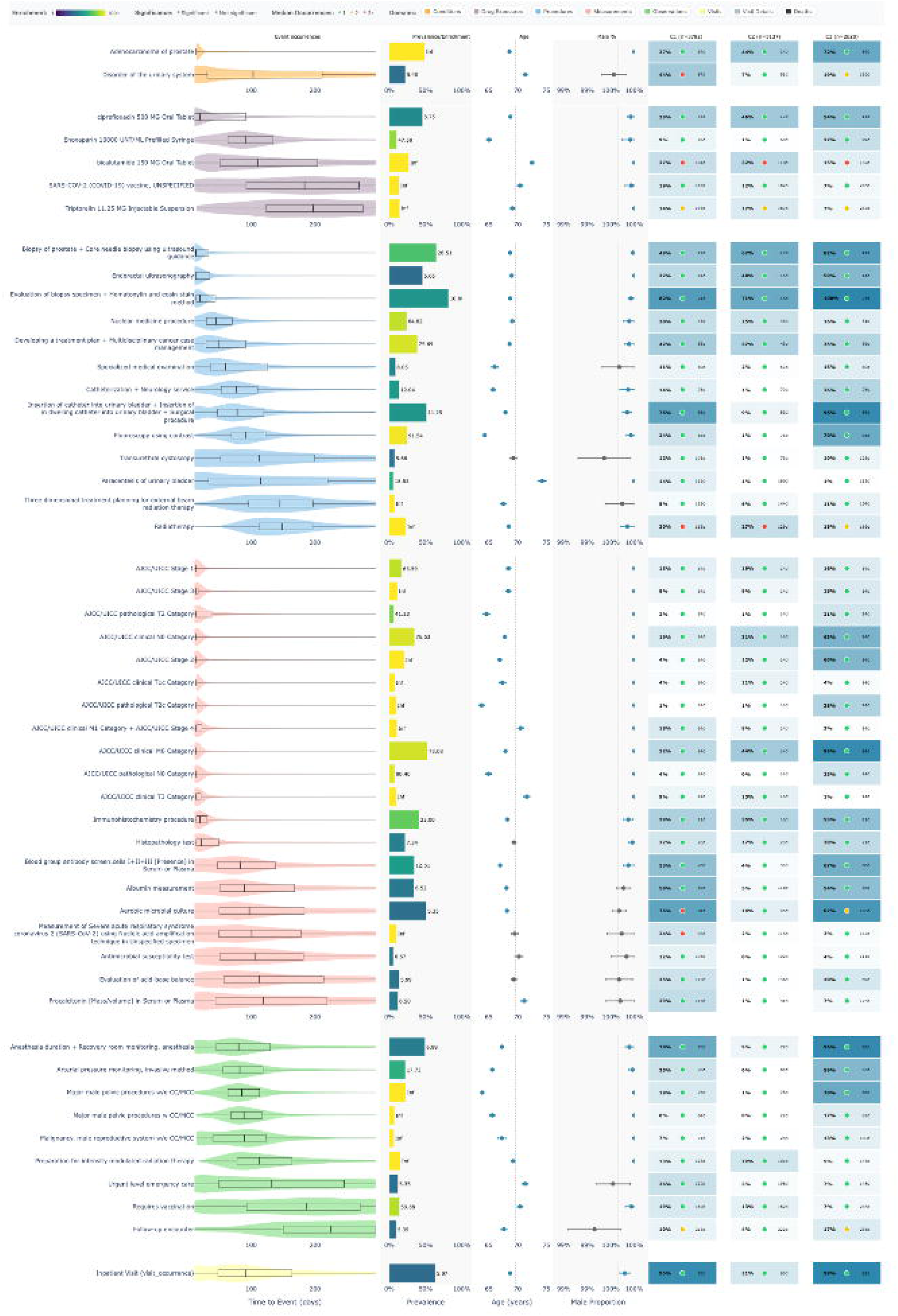
Clustering of prostate cancer patients. The heatmap shows the 50 concepts with the highest between-cluster variance. Event-occurrence plots show concept timing across the observation period. The prevalence/enrichment panel summarizes prevalence in the target cohort and enrichment versus controls (Inf indicates no control occurrences). The age/male panel shows distributions and confidence intervals relative to the average. The cluster panel shows cluster-specific prevalence and median occurrence counts.

Downstream care patterns further separated the phenotypes (Figure 4). C3 showed the strongest peri-procedural/inpatient signature consistent with definitive local therapy, with high inpatient visits (99.4%), anesthesia duration and recovery room monitoring (96.4%), major male pelvic procedures w/o CC/MCC (70.5%), urinary bladder catheter insertion/indwelling catheter procedure (95.9%) and invasive arterial pressure monitoring (59.5%). C2 showed an outpatient definitive management pattern enriched for radiotherapy and androgen deprivation therapy related concepts, including radiotherapy (27.4%), preparation for intensity modulated radiation therapy (21.8%), bicalutamide (32.4%) and triptorelin (17.2%). In contrast, C1 exhibited a higher-complexity/acuity-enriched course with greater genitourinary complication and acute-care signals, including disorder of the urinary system (43.5%), urgent level emergency care (25.8%), paracentesis of urinary bladder (13.7%), procalcitonin (29.0%) and antimicrobial susceptibility testing (12.4%), supporting a complication/acute-utilization trajectory during the first year after diagnosis. (Appendix 6)

The validators rated 466 diagnosis–concept pairs (296 lung, 170 prostate) as *direct*, *indirect*, or *noise* (Appendices 7 and 8). Overall 31.3% of extracted concepts were labeled as *direct* with 57.5% as *indirect* and 11.2% as *noise*. All raters labeled most pairs as *indirect* (MD1: 55.6%; MD2: 50.4%; LLM: 66.3%). MD2 assigned the highest proportion of *direct* labels overall (37.6%) compared with MD1 (29.8%) and the LLM (26.6%), while *noise* labels were relatively infrequent across raters (MD1: 14.6%; MD2: 12.0%; LLM: 7.1%). Similar patterns were observed within the lung and prostate subsets, with the LLM consistently assigning the lowest proportion of *noise* labels (7.1% lung; 7.0% prostate) (Table 2).

**Table 2.**
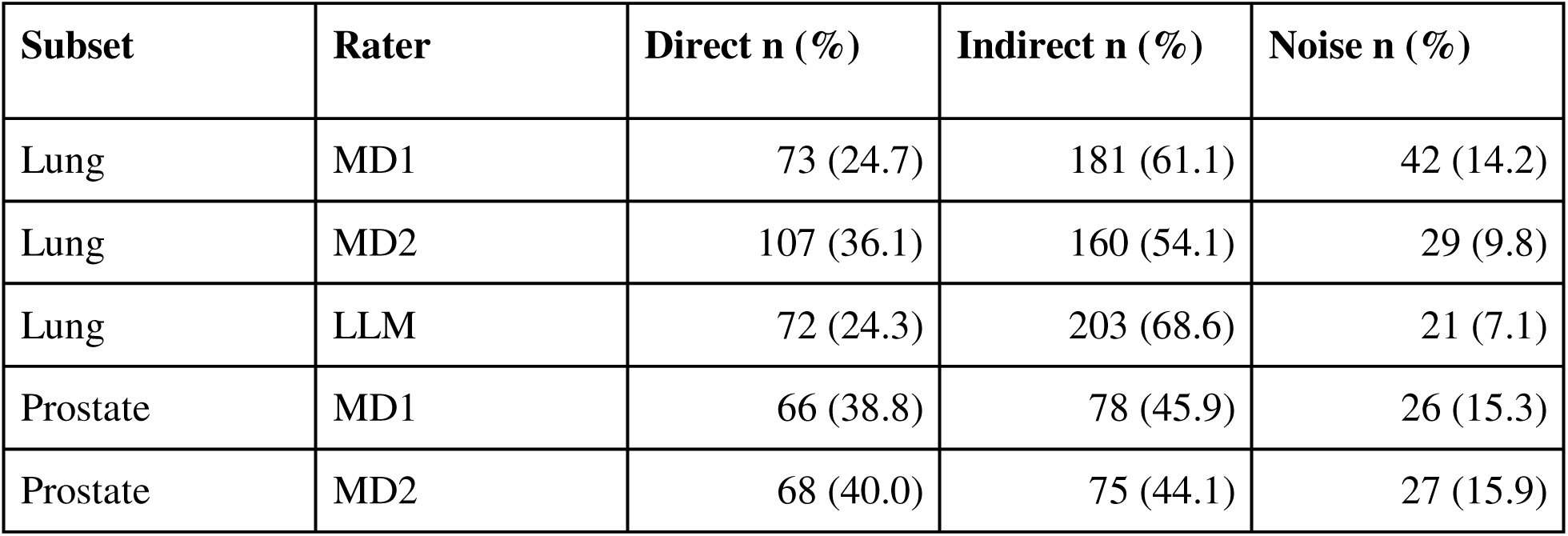
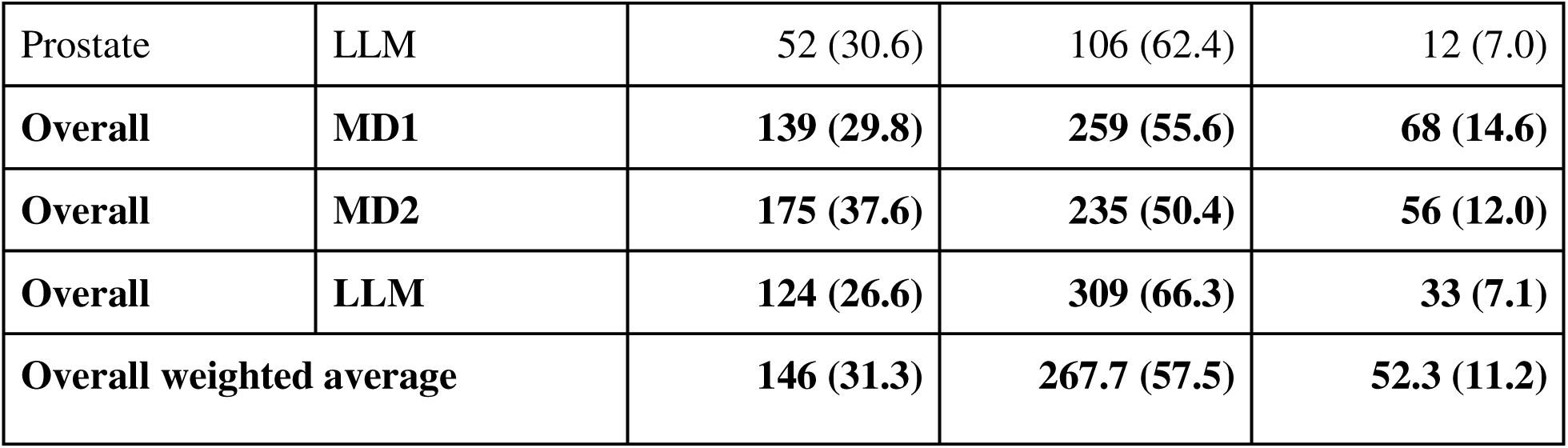
Extracted concept label distribution by rater across lung, prostate and overall subsets.

| Subset | Rater | Direct n (%) | Indirect n (%) | Noise n (%) |
| --- | --- | --- | --- | --- |
| Lung | MD1 | 73 (24.7) | 181 (61.1) | 42 (14.2) |
| Lung | MD2 | 107 (36.1) | 160 (54.1) | 29 (9.8) |
| Lung | LLM | 72 (24.3) | 203 (68.6) | 21 (7.1) |
| Prostate | MD1 | 66 (38.8) | 78 (45.9) | 26 (15.3) |
| Prostate | MD2 | 68 (40.0) | 75 (44.1) | 27 (15.9) |
| Prostate | LLM | 52 (30.6) | 106 (62.4) | 12 (7.0) |
| **Overall** | **MD1** | **139 (29.8)** | **259 (55.6)** | **68 (14.6)** |
| **Overall** | **MD2** | **175 (37.6)** | **235 (50.4)** | **56 (12.0)** |
| **Overall** | **LLM** | **124 (26.6)** | **309 (66.3)** | **33 (7.1)** |
| **Overall weighted average** |  | **146 (31.3)** | **267.7 (57.5)** | **52.3 (11.2)** |

Of the concepts excluded by the workflow, the LLM labeled 4.0% (n=437) as *direct*, 33.2% (n=3,612) as *indirect*, and 62.8% (n=6,827) as *noise* (Table 3). Excluded *direct* concepts were rare (median 2 patients) but showed high enrichment, with *Inf* indicating zero control counts.

**Table 3.** Disregarded concept label distribution across lung, prostate and overall subsets.

| Subset | Label | n (%) | Patients (q1/q2/q3) | Enrichment (q1/q2/q3) |
| --- | --- | --- | --- | --- |
| Lung | Direct | 280 (5.2) | 1/2/4 | 3.65/Inf*/Inf* |
| Lung | Indirect | 1724 (32.2) | 1/4/16 | 0.65/1.18/6.38 |
| Lung | Noise | 3353 (62.6) | 1/2/5 | 0.30/0.91/2.73 |
| Prostate | Direct | 156 (2.8) | 1/3/5 | 2.37/Inf*/Inf* |
| Prostate | Indirect | 1889 (34.2) | 2/5/29 | 0.92/1.40/7.03 |
| Prostate | Noise | 3475 (63.0) | 1/2/8 | 0.62/1.13/Inf* |
| **Overall** | **Direct** | **436 (4.0)** | **1/2/4** | **2.77/Inf\*/Inf\*** |
| **Overall** | **Indirect** | **3613 (33.2)** | **1/4/21** | **0.80/1.32/6.46** |
| **Overall** | **Noise** | **6828 (62.8)** | **1/2/7** | **0.46/0.92/4.56** |

\*: Enrichment is marked as Inf if the denominator (control cohort count) is zero.

Overall, agreement was highest between MD1 and the LLM (79.0%, _κ_ = 0.612, 95% CI 0.546–0.657) and lowest between MD2 and the LLM (68.7%, _κ_ = 0.438, 95% CI 0.368–0.517). Agreement between MD1 and MD2 was 70.8% (_κ_ = 0.505, 95% CI 0.437–0.571). Similar patterns were observed in the lung and prostate subsets (Appendix 9).

## DISCUSSION

Existing trajectory methods often depend on predefined states or constrained, manually curated concept sets.[10,36,37] Our earlier OMOP trajectory tools automate sequence construction from observed events,[9] but not the upstream selection of cohort-relevant concepts. CohortContrast addresses that step by prioritizing and aggregating concepts across a broad OMOP-compatible concept space before downstream trajectory analysis. Within OHDSI, FeatureExtraction can generate a similarly broad initial candidate universe at the level of Phase I when configured to return all eligible covariates, but it does not perform the subsequent enrichment, temporal filtering, and aggregation steps that condense that universe into an interpretable cohort-specific concept set.[38]

Rather than replacing downstream trajectory methods, CohortContrast addresses the missing upstream step of converting a broad OMOP-derived candidate concept space into a cohort-specific, analysis-ready concept set. Its contribution therefore lies in reproducible prioritization, temporal specificity filtering, and semantic aggregation, enabling downstream trajectory analyses to operate on a markedly reduced yet clinically interpretable representation.

Although Phase III uses the same enrichment-testing framework as Phase II, it provides an important additional filtering step by reducing temporal and global background noise. In the lung cancer cohort, this phase removed non-specific concepts such as chronic disease monitoring, screening programs and COVID-19 vaccination related pathways, which were likely enriched because the selected observation windows overlapped broader healthcare system effects rather than reflecting lung cancer–specific treatment patterns.

At the same time, Phase III highlights an important dependence on the composition of the underlying OMOP instance. In the prostate cancer cohort, the same filtering step also removed clinically plausible concepts (eg, chemotherapy, endoscopic biopsy, ultrasonography and cytologic testing) because these concepts were relatively prevalent in the matched background population. In our database, which is restricted to patients with cancer diagnoses, some oncology-related concepts are common at baseline and therefore not reported.

Across both cancer case studies, clustering of CohortContrast-derived concept sets yielded three clinically interpretable, exploratory trajectory archetypes. In lung cancer, clusters broadly reflected (i) advanced/metastatic actively treated pathways, (ii) rapid-decline/limited-treatment pathways, and (iii) earlier-stage procedure-heavy pathways, aligned with heterogeneity described in prior administrative claims pathway analyses.[39] In prostate cancer, clusters reflected (i) older higher-complexity courses with urinary/acute-care signals, (ii) outpatient definitive management enriched for radiotherapy and androgen-deprivation therapy, and (iii) younger peri-procedural definitive local treatment patterns, similar to prior national claims clustering results.[40]

Clustering treatment trajectories directly from real-world data is inherently challenging because care pathways are heterogeneous, temporally irregular, and influenced by factors beyond disease biology.[41] Even within cancer, where treatment regimens are comparatively well defined, patients may switch, pause or discontinue therapies for many reasons and the same clinical concepts can appear with substantial variability in timing, frequency and sequencing across individuals. In this context, the fact that CohortContrast yields clinically interpretable trajectory archetypes in a largely unsupervised, upstream manner is notable: by prioritizing cohort-relevant concepts and reducing high-dimensional noise before clustering, the workflow mitigates some of the fragmentation that typically obscures structure in RWD-derived trajectories.

The validation results support CohortContrast as a practical concept-reduction workflow that retains clinically relevant signals while minimizing residual noise. Clinician agreement on extracted diagnosis–concept pairs was moderate (MD1–MD2 _κ_ = 0.505), highlighting that direct-versus-indirect labeling is inherently interpretive. The LLM showed comparable concordance to clinician–clinician agreement (MD1–LLM _κ_ = 0.612; MD2–LLM _κ_ = 0.438), supporting its use as a scalable proxy rater for initial triage rather than exhaustive clinician review. Only 7.1% of extracted pairs were labeled as noise by the LLM, and among all excluded concepts (10,877), only 4.0% were labeled as direct. These excluded concepts labeled as direct were typically very rare, with a median of two patients, so they were preferentially removed during statistical filtering and were less likely to be retained through hierarchy-based aggregation when no retained parent concept was available. In practice, many would also fall below small-cell thresholds for privacy-preserving result sharing, further limiting their analytic usefulness.

Beyond reducing the concept space, CohortContrast increased information density. Using LLM-based categorization of concept interpretability, the proportion of concepts labeled *direct* increased 4.1-fold in lung and 8.5-fold in prostate, indicating substantial enrichment for interpretable cohort-discriminative concepts (Tables 2 and 3).

Across both case studies, CohortContrast automated cohort characterization in the OMOP CDM by reducing concept dimensionality, prioritizing cohort-relevant signals, and organizing them into interpretable groupings that translate into clinically meaningful summaries and analysis-ready artifacts. In the lung cancer case study, the workflow recovered concepts aligned with established clinical constructs, including stage-related patterns (eg, cM1/Stage 4 vs cM0/Stage 1A/pN0) and distinct treatment-phase signatures (eg, systemic therapy and radiotherapy planning with supportive medications versus procedure-heavy respiratory/pleural care). These outputs are directly useful in real-world evidence studies: they can define clinically interpretable subgroups and treatment pathways for HTA-style analyses, supply cohort-specific covariates for adjustment, matching, or stratification, and be rerun under alternative exposures, index definitions, or time-at-risk windows to identify enriched complications for signal detection, safety evaluation, and downstream trajectory analyses.

Beyond analytic utility, CohortContrast was implemented to support practical and shareable cohort exploration. The package can generate either patient-level results for detailed local analysis or privacy-preserving summary outputs with configurable small-cell suppression, allowing concept-level summaries, clustering structure, overlap patterns, and occurrence ordering to be reviewed without transferring row-level data. These outputs can be delivered through the GUI, including web-server deployment, to provide an accessible descriptive overview of cohorts of interest while supporting reproducibility and reuse through the publicly available code and released results.[21,22]

## Limitations

The first limitation is the potential loss of clinical specificity during hierarchical concept aggregation. While mapping concepts to higher-level categories improves interpretability and reduces redundancy, it can also obscure important distinctions. For example, “Administration of medication” may subsume more specific interventions such as “Chemotherapy”. To mitigate this, we preserved child concepts when their prevalence exceeded that of the parent, retaining high-frequency specific concepts that might otherwise be lost.

The second limitation concerns vocabulary hierarchy–based drug aggregation, which may underperform when medications are recorded at the package or product level. In such cases, similar drug concepts may not be grouped together if their shared RxNorm parent concept is not present in the retained set. This can fragment related drug concepts, reduce apparent frequency and increase the likelihood of exclusion during filtering.

A further limitation is the modest geometric separation of the patient clusters, as reflected by low silhouette scores (0.143 in lung cancer and 0.102 in prostate cancer). Although the clusters were clinically interpretable, these values indicate overlapping subgroup structure, which is expected in high-dimensional, temporally derived medical concept spaces. Similar trajectory-oriented medical clustering studies have reported modest silhouette values while still identifying clinically meaningful patterns, and standard silhouette has recognized limitations in sparse longitudinal clinical data.[41–43]

## CONCLUSION

CohortContrast is an OMOP-compatible R package for enrichment-based identification and aggregation of cohort-relevant clinical concepts for downstream treatment trajectory analysis. In lung and prostate cancer case studies, the workflow reduced concept dimensionality substantially while preserving clinically interpretable patterns and identifying three exploratory patient subgroups in both cohorts. Across raters, most extracted diagnosis-concept pairs were judged direct or indirect rather than noise. These findings support CohortContrast as a scalable and reproducible workflow for hypothesis generation, cohort refinement, and exploratory real-world evidence research in OMOP CDM datasets.

## AUTHOR CONTRIBUTIONS

All authors participated in the revision process and have approved the submitted version. The work reported in the paper has been performed by the authors, unless clearly specified in the text. *Conceptualization* – Markus Haug, Raivo Kolde. *Data curation* – Markus Haug, Raivo Kolde, Marek Oja, Sulev Reisberg, Jaak Vilo. Formal Analysis – Markus Haug, Raivo Kolde. *Funding acquisition* – Raivo Kolde. Methodology – Markus Haug, Raivo Kolde, Kerli Mooses, Sirli Tamm, Helene Loorents. Project administration – Markus Haug, Raivo Kolde. Resources – Markus Haug, Raivo Kolde. Software – Markus Haug, Raivo Kolde. Supervision – Raivo Kolde. Validation – Markus Haug, Neeme Ilves, Nikita Umov, Hendrik Šuvalov. Visualization – Markus Haug, Raivo Kolde. Writing – original draft – Markus Haug. Writing – Markus Haug, Raivo Kolde, Hendrik Šuvalov.

## DATA AVAILABILITY STATEMENT

There are legal restrictions on sharing de-identified data. According to legislative regulation and data protection law in Estonia, the authors cannot publicly release the data received from the health data registers in Estonia.

An interactive browser of the cell-suppressed, aggregate CohortContrast results (excluding the validation outputs) is available at: http://omop-apps.cloud.ut.ee/CohortContrast/ (accessed April 21, 2026).

## ETHICS STATEMENT

The study was approved by the Research Ethics Committee of the University of Tartu (300/T-23 and No. 330/T-10) and the Estonian Committee on Bioethics and Human Research (1.1-12/653) and the requirement for informed consent was waived.

## Abbreviations

CDM: common data model
CHI2Y: Chi-squared test for two proportions with Yates continuity correction
CRAN: Comprehensive R Archive Network
GUI: graphical user interface
LLM: large language model
LRTEST: Univariate logistic regression’s Wald’s test
MD: medical doctor
OHDSI: Observational Health Data Sciences and Informatics
OMOP: Observational Medical Outcomes Partnership
PCA: principal component analysis
RWD: real-world data

## Article category

Research and Applications

## Novelty and Impact

We introduce CohortContrast, an open-source OMOP-compatible R package for enrichment-based cohort contrast and automated aggregation of cohort-relevant clinical concepts to support exploratory treatment pattern analysis. In lung (n=7,579) and prostate (n=11,547) cancer cohorts, the workflow identified clinically relevant concepts across diagnostic, therapeutic, monitoring and supportive-care domains while substantially reducing concept dimensionality. CohortContrast enables data-driven cohort characterization and provides a foundation for downstream trajectory and process-oriented analyses.

