## Supplementary figures and images for "CohortContrast: An R Package for Enrichment-Based Identification of Clinically Relevant Concepts in OMOP CDM Data"

### Appendix 1

**Appendix 1: In-depth workflow for CohortContrast**

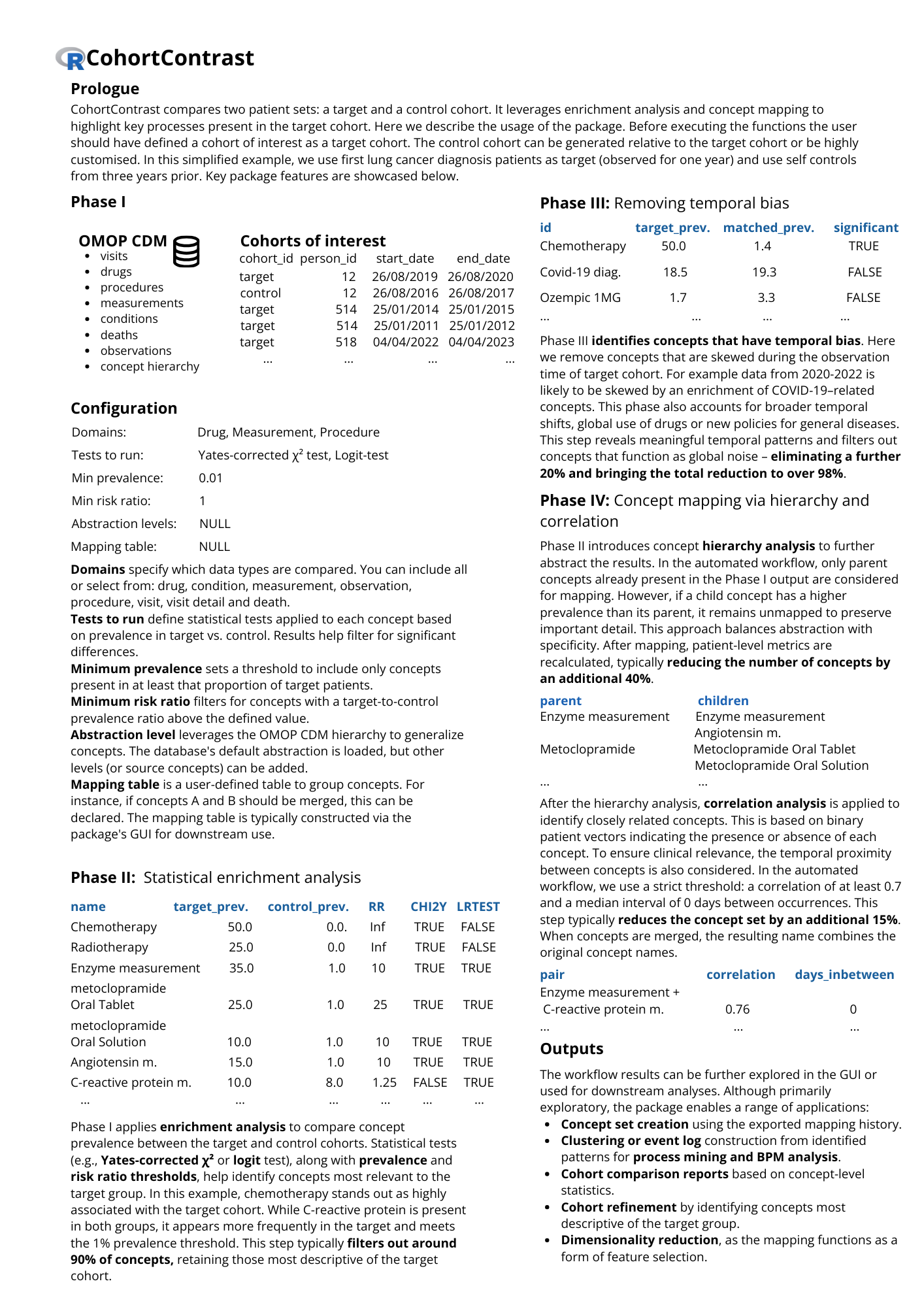

### Appendix 2

**Appendix 2: Clustering workflow**

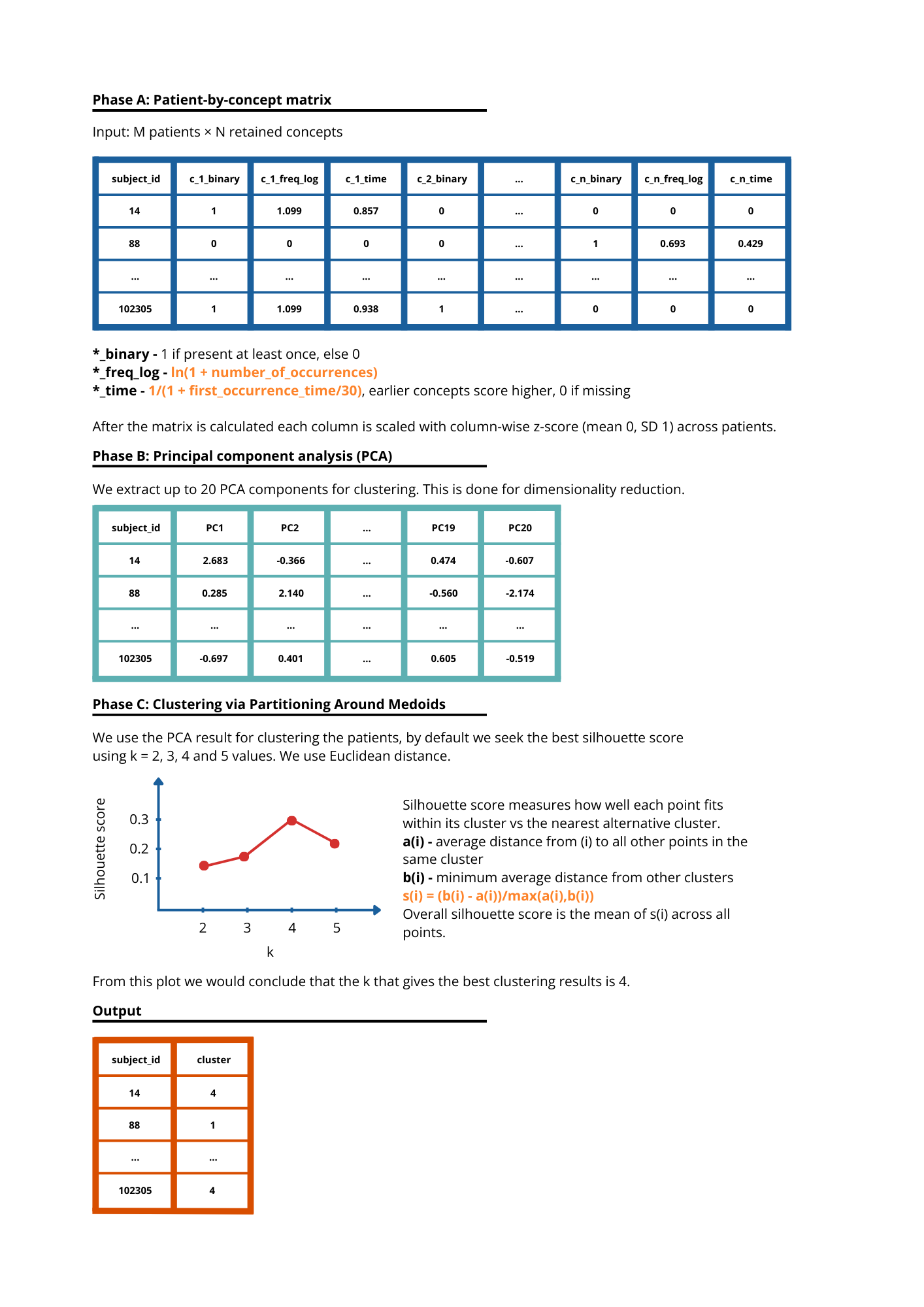
