## Appendix 3 for "CohortContrast: An R Package for Enrichment-Based Identification of Clinically Relevant Concepts in OMOP CDM Data"

**Appendix 3: Demographics for the observed target cohorts**

| **Cohort** | **Lung cancer** | **Prostate cancer** |
| --- | --- | --- |
| **Patient count** | 7579 | 11 547 |
| **Male %** | 69.05% | 99.66% |
| **Age (25th pct)** | 63.87 | 63.80 |
| **Age (median)** | 70.64 | 69.95 |
| **Age (75th pct)** | 77.89 | 76.39 |
| **Follow-up time (25th pct)** | 3.14 months | 12.01 months |
| **Follow-up time (median)** | 10.3 months | 12.04 months |
| **Follow-up time (75th pct)** | 12.04 months | 12.04 months |
| **Year of diagnosis (25th pct)** | 2016 | 2016 |
| **Year of diagnosis (median)** | 2018 | 2018 |
| **Year of diagnosis (75th pct)** | 2020 | 2020 |
| **Condition recorded for** | 100% | 100% |
| **Procedure recorded for** | 99.5% | 98.7% |
| **Measurement recorded for** | 99.4% | 97.9% |
| **Drug recorded for** | 94.6% | 97.3% |
| **Observation recorded for** | 99.9% | 99.9% |
| **Visit recorded for** | 99.8% | 99.8% |
| **Visit detail recorded for** | 99.9% | 99.9% |
| **Death recorded for** | 47.87% | 5.80% |
| **Median death time in cohort** | 99 days | 149 days |

**Note: Both cohorts’ index date was shifted -14 days for detecting concepts related to pre-diagnosis**
