## Appendix 4 for "CohortContrast: An R Package for Enrichment-Based Identification of Clinically Relevant Concepts in OMOP CDM Data"

**Appendix 4: Instructions for medical doctors and the LLM**

**Instructions**

You as a medical doctor are validating whether listed clinical concepts are meaningfully related to a given medical condition.

Context for concept extraction:

These concepts were statistically extracted by comparing patient data from two time periods:

- Target cohort: 1 year following the patient's first diagnosis of the condition

- Control cohort: 1 year period three years before their first diagnosis

Concepts with statistically significant enrichment in the target period were extracted for your assessment.

Example of a concepts (say we are looking at a condition X):

CONCEPT_NAME | HERITAGE

Pneumonia	| condition_occurrence

Claim	| death

This means that for condition X, the patients got elevated (target period vs control period) diagnosis of pneumonia and death claims (died within 1 year).

Relationship labels:

1 (Direct relationship)

Use 1 when the concept would commonly be considered closely tied to the condition in typical clinical practice, such as:

- The condition itself, clear synonym, or subtype

- A hallmark/pathognomonic feature strongly suggestive of the condition

- A diagnostic procedure/test commonly used to confirm/stage/evaluate this condition (especially if organ- or condition-specific)

- A standard treatment strongly associated with management of this condition (including common definitive therapies in routine care)

0 (Indirect relationship)

Use 0 when the concept is clinically plausible in the condition’s context but not specific, such as:

- Common symptoms/signs, broad labs, generic imaging or procedures

- Typical comorbidities, complications, or supportive care that may co-occur but are not uniquely indicative

- Generic care utilization (visits/admissions), monitoring, or administrative processes that could increase after diagnosis

- Drugs/therapies used for many conditions without a strong, condition-specific linkage

-1 (Noise / unrelated)

Use -1 when the concept is not meaningfully related in typical practice:

- Clearly unrelated system/context with no plausible clinical link

- Too vague to interpret and not plausibly connected

- Likely to occur similarly across many unrelated conditions without a reasonable link

**Prompt** (pruned example)

Condition: Malignant neoplasm of lung

Nausea and vomiting | condition_occurrence

Dysphagia | condition_occurrence

etoposide 100 MG Oral Capsule | drug_exposure

**Response schema**

{ "classifications": [ {

"concept_name": "<CONCEPT_NAME>",

"relationship": 1 | 0 | -1,

"rationale": "<concise clinical justification in ≤10 words>"

} ]}
