## Appendix 5 for "CohortContrast: An R Package for Enrichment-Based Identification of Clinically Relevant Concepts in OMOP CDM Data"

**Appendix 5: Top 50 concepts with the highest between-cluster variance for the lung cancer cohort**

| **Concept Name** | **C1 Prevalence (%)** | **C1 Median Time** | **C2 Prevalence (%)** | **C2 Median Time** | **C3 Prevalence (%)** | **C3 Median Time** |
| --- | --- | --- | --- | --- | --- | --- |
| Death Certificate | 67.04 | 124.0 | 55.51 | 70.0 | 10.23 | 183.0 |
| acetaminophen 500 MG / codeine phosphate 30 MG Oral Tablet | 45.95 | 109.0 | 23.00 | 112.0 | 33.63 | 114.0 |
| dexamethasone 4 MG Oral Tablet | 27.26 | 126.0 | 5.35 | 61.5 | 4.08 | 237.5 |
| Bilirubin measurement | 87.62 | 102.0 | 46.42 | 111.0 | 87.95 | 99.0 |
| Cytologic test | 77.28 | 15.0 | 45.69 | 15.0 | 54.62 | 15.0 |
| Immunohistochemistry procedure | 75.92 | 18.0 | 32.16 | 15.0 | 69.91 | 19.5 |
| Blood group antibody screen.cells I+II+III [Presence] in Serum or Plasma | 59.92 | 49.0 | 23.69 | 33.0 | 90.41 | 33.0 |
| Albumin measurement | 62.94 | 64.0 | 17.22 | 48.0 | 73.60 | 49.0 |
| Histopathology test | 57.04 | 17.0 | 28.58 | 15.0 | 49.85 | 22.0 |
| Procalcitonin [Mass/volume] in Serum or Plasma | 61.89 | 78.0 | 19.88 | 37.0 | 41.99 | 66.0 |
| Evaluation of acid-base balance | 49.44 | 50.0 | 17.64 | 21.0 | 55.60 | 54.0 |
| Spirometry | 27.94 | 15.0 | 24.81 | 16.0 | 58.60 | 22.0 |
| AJCC/UICC clinical M0 Category | 19.13 | 14.0 | 24.23 | 14.0 | 63.62 | 14.0 |
| Peripheral blood smear examination, light microscopy | 45.24 | 90.0 | 18.14 | 41.0 | 28.37 | 106.0 |
| AJCC/UICC clinical M1 Category + AJCC/UICC Stage 4 | 42.96 | 14.0 | 34.28 | 14.0 | 8.26 | 14.0 |
| Diffusion capacity of lung | 19.23 | 15.0 | 13.91 | 17.0 | 54.82 | 18.0 |
| HIV 1+2 Ab+HIV1 p24 Ag [Presence] in Serum or Plasma by Immunoassay | 18.14 | 24.0 | 8.78 | 18.0 | 42.08 | 22.0 |
| Polymerase chain reaction analysis | 33.71 | 46.0 | 8.17 | 19.0 | 18.14 | 54.0 |
| AJCC/UICC clinical N2 Category | 25.16 | 14.0 | 20.34 | 14.0 | 4.92 | 14.0 |
| Red blood cell test | 27.37 | 93.5 | 4.89 | 72.0 | 17.11 | 107.5 |
| Measurement of Severe acute respiratory syndrome coronavirus 2 (SARS-CoV-2) using Nucleic acid amplification technique in Unspecified specimen | 27.81 | 61.0 | 4.31 | 33.0 | 15.14 | 51.0 |
| AJCC/UICC clinical T4 Category | 23.70 | 14.0 | 13.37 | 14.0 | 2.46 | 14.0 |
| AJCC/UICC pathological N0 Category | 0.24 | 14.0 | 1.46 | 14.0 | 36.48 | 14.0 |
| AJCC/UICC Stage 1A | 0.88 | 14.0 | 4.31 | 14.0 | 23.70 | 14.0 |
| Death | 66.53 | 124.0 | 54.93 | 71.0 | 10.03 | 185.0 |
| Urgent level emergency care | 44.15 | 67.0 | 8.86 | 20.5 | 20.30 | 130.0 |
| Died in hospital | 25.43 | 121.0 | 29.28 | 62.0 | 5.51 | 171.0 |
| Arterial pressure monitoring, invasive method | 10.34 | 51.0 | 3.43 | 14.0 | 57.52 | 31.0 |
| Evaluation of biopsy specimen | 79.38 | 17.0 | 43.03 | 15.0 | 97.89 | 23.0 |
| Developing a treatment plan + Multidisciplinary cancer case management | 81.89 | 28.0 | 50.62 | 24.0 | 78.96 | 34.0 |
| Bronchoscopy | 76.03 | 14.0 | 42.60 | 14.0 | 50.64 | 25.0 |
| Hematoxylin and eosin stain method | 76.13 | 17.0 | 37.37 | 15.0 | 43.95 | 18.0 |
| Chemotherapy | 62.39 | 104.0 | 30.12 | 104.0 | 40.71 | 114.0 |
| Endoscopic biopsy | 61.28 | 14.0 | 30.39 | 14.0 | 29.01 | 14.0 |
| Analysis using real time PCR + Chromosome analysis, cytogenetic procedure AND/OR molecular  biology method | 59.99 | 38.0 | 16.06 | 21.0 | 34.96 | 49.0 |
| Insertion of tracheostomy tube + Surgical procedure | 22.21 | 75.0 | 10.79 | 19.0 | 94.84 | 28.0 |
| Mycobacterial microscopy | 43.54 | 15.0 | 22.00 | 15.0 | 28.22 | 14.0 |
| Drainage of pleural cavity | 19.36 | 40.0 | 6.74 | 15.0 | 72.42 | 29.0 |
| Radiotherapy + Three dimensional treatment planning for external beam radia- tion therapy | 36.05 | 111.0 | 20.88 | 79.0 | 15.19 | 140.0 |
| Insertion of catheter into urinary bladder + Insertion of indwelling catheter into urinary bladder | 24.04 | 74.0 | 11.52 | 22.0 | 44.49 | 50.0 |
| Plain radiography | 14.85 | 56.0 | 5.01 | 21.0 | 33.78 | 42.0 |
| Endoscopy of trachea | 2.17 | 31.0 | 2.77 | 14.0 | 52.75 | 31.0 |
| Thoracentesis | 11.02 | 29.0 | 3.70 | 20.0 | 35.40 | 36.0 |
| Administration of medication | 16.41 | 112.0 | 2.20 | 162.5 | 25.12 | 47.0 |
| Catheterization | 7.87 | 64.0 | 2.23 | 21.0 | 36.04 | 43.0 |
| Specialized medical examination | 2.00 | 33.0 | 2.08 | 14.0 | 25.61 | 21.0 |
| Intubation | 0.71 | 37.5 | 1.27 | 20.0 | 19.76 | 29.0 |
| Neurology service | 0.44 | 68.0 | 0.27 | 19.0 | 18.53 | 49.0 |
| Inpatient Visit | 98.20 | 53.0 | 72.61 | 19.0 | 99.95 | 43.0 |
| Inpatient Visit (visit_occurrence) | 96.95 | 67.0 | 67.72 | 42.0 | 99.85 | 48.0 |
