## Appendix 6 for "CohortContrast: An R Package for Enrichment-Based Identification of Clinically Relevant Concepts in OMOP CDM Data"

**Appendix 6: Top 50 concepts with the highest between-cluster variance for the prostate cancer cohort**

| **Concept Name** | **C1 Prevalence (%)** | **C1 Median Time** | **C2 Prevalence (%)** | **C2 Median Time** | **C3 Prevalence (%)** | **C3 Median Time** |
| --- | --- | --- | --- | --- | --- | --- |
| Adenocarcinoma of prostate | 37.07 | 14.0 | 44.01 | 14.0 | 72.15 | 14.0 |
| Disorder of the urinary system | 43.52 | 97.0 | 6.99 | 81.0 | 19.37 | 126.0 |
| ciprofloxacin 500 MG Oral Tablet | 38.87 | 44.0 | 45.61 | 17.0 | 54.30 | 14.0 |
| bicalutamide 150 MG Oral Tablet | 26.89 | 104.0 | 32.39 | 109.5 | 14.73 | 131.5 |
| Triptorelin 11.25 MG Injectable Suspension | 14.23 | 195.0 | 17.19 | 187.0 | 7.46 | 237.0 |
| SARS-COV-2 (COVID-19) vaccine, UNSPECIFIED | 18.61 | 163.0 | 12.28 | 182.0 | 6.93 | 248.0 |
| Enoxaparin 10000 UNT/ML Prefilled Syringe | 9.44 | 78.0 | 1.27 | 67.5 | 27.09 | 99.0 |
| AJCC/UICC clinical M0 Category | 32.28 | 14.0 | 44.11 | 14.0 | 96.27 | 14.0 |
| Aerobic microbial culture | 75.57 | 94.0 | 13.72 | 95.0 | 86.68 | 100.0 |
| Immunohistochemistry procedure | 39.42 | 21.0 | 35.22 | 16.0 | 52.66 | 21.0 |
| AJCC/UICC clinical N0 Category | 18.85 | 14.0 | 31.48 | 14.0 | 62.79 | 14.0 |
| Blood group antibody screen.cells I+II+III [Presence] in Serum or Plasma | 53.07 | 78.0 | 3.76 | 94.0 | 66.51 | 88.0 |
| Albumin measurement | 57.85 | 89.0 | 5.49 | 118.0 | 53.65 | 88.0 |
| Histopathology test | 21.66 | 26.0 | 16.66 | 20.0 | 30.37 | 21.0 |
| AJCC/UICC Stage 2 | 4.12 | 14.0 | 12.30 | 14.0 | 60.16 | 14.0 |
| Evaluation of acid-base balance | 24.56 | 120.0 | 1.44 | 136.5 | 20.09 | 89.0 |
| Procalcitonin [Mass/volume] in Serum or Plasma | 28.98 | 118.0 | 1.09 | 91.0 | 6.54 | 125.0 |
| AJCC/UICC Stage 3 | 8.33 | 14.0 | 8.58 | 14.0 | 18.30 | 14.0 |
| AJCC/UICC clinical M1 Category + AJCC/UICC Stage 4 | 18.35 | 14.0 | 8.25 | 14.0 | 1.90 | 14.0 |
| Measurement of Severe acute respiratory syndrome coronavirus 2 (SARS-CoV-2) using Nucleic acid amplification technique in Unspecified specimen | 23.98 | 95.0 | 2.02 | 115.0 | 3.46 | 142.0 |
| AJCC/UICC pathological T2c Category | 1.11 | 14.0 | 0.58 | 14.0 | 37.98 | 14.0 |
| AJCC/UICC clinical T3 Category | 9.47 | 14.0 | 13.08 | 14.0 | 1.14 | 14.0 |
| AJCC/UICC pathological N0 Category | 4.34 | 14.0 | 0.35 | 14.0 | 22.91 | 14.0 |
| AJCC/UICC pathological T2 Category | 1.72 | 14.0 | 0.84 | 14.0 | 21.39 | 14.0 |
| Antimicrobial susceptibility test | 12.35 | 106.0 | 0.25 | 102.0 | 4.30 | 111.0 |
| AJCC/UICC pathological T3a Category | 3.78 | 14.0 | 0.37 | 14.0 | 14.12 | 14.0 |
| Anesthesia duration + Recovery room monitoring, anesthesia | 70.12 | 70.0 | 8.90 | 72.0 | 96.35 | 86.0 |
| Major male pelvic procedures w/o CC/MCC | 18.67 | 75.5 | 0.82 | 78.0 | 70.47 | 88.0 |
| Arterial pressure monitoring, invasive method | 24.56 | 78.0 | 0.47 | 84.0 | 59.47 | 83.0 |
| Preparation for intensity modulated radiation therapy | 12.74 | 125.0 | 21.80 | 101.0 | 4.98 | 143.0 |
| Requires vaccination | 18.96 | 169.0 | 12.67 | 182.0 | 7.19 | 246.0 |
| Urgent level emergency care | 25.83 | 120.0 | 3.33 | 196.5 | 6.58 | 147.5 |
| Follow-up encounter | 10.47 | 213.0 | 4.40 | 222.0 | 16.70 | 238.0 |
| Major male pelvic procedures w CC/MCC | 8.20 | 84.0 | 0.10 | 22.0 | 17.35 | 92.0 |
| Malignancy, male reproductive system w/o CC/MCC | 7.24 | 71.0 | 1.54 | 27.5 | 12.82 | 100.0 |
| Malignancy, male reproductive system w CC | 11.87 | 49.0 | 0.82 | 25.0 | 3.27 | 108.0 |
| Urinary bladder stoma present | 11.92 | 145.0 | 0.93 | 127.5 | 0.42 | 121.5 |
| Evaluation of biopsy specimen + Hematoxylin and eosin stain method | 81.84 | 21.0 | 71.07 | 16.0 | 99.96 | 23.0 |
| Biopsy of prostate + Core needle biopsy using ultrasound guidance | 48.70 | 14.0 | 67.04 | 14.0 | 81.96 | 14.0 |
| Insertion of catheter into urinary bladder + Insertion of in dwelling catheter into urinary bladder + Surgical procedure | 76.34 | 68.0 | 9.09 | 82.5 | 95.93 | 84.0 |
| Endorectal ultrasonography | 32.10 | 14.0 | 48.39 | 14.0 | 58.98 | 14.0 |
| Fluoroscopy using contrast | 23.98 | 85.0 | 0.58 | 75.0 | 70.02 | 95.0 |
| Nuclear medicine procedure | 27.58 | 45.0 | 24.82 | 43.0 | 16.02 | 51.0 |
| Radiotherapy | 20.44 | 163.0 | 27.39 | 129.0 | 17.62 | 186.0 |
| Catheterization + Neurology service | 14.28 | 76.0 | 0.95 | 69.5 | 35.65 | 78.0 |
| Specialized medical examination | 10.76 | 60.0 | 2.47 | 57.0 | 15.18 | 60.0 |
| Transurethral cystoscopy | 11.77 | 105.0 | 1.36 | 73.0 | 10.16 | 123.0 |
| Paracentesis of urinary bladder | 13.70 | 111.0 | 0.86 | 160.0 | 0.76 | 112.0 |
| Change of cystostomy tube | 12.37 | 167.0 | 1.15 | 161.0 | 0.46 | 122.0 |
| Inpatient Visit (visit_occurrence) | 95.11 | 88.0 | 20.79 | 98.0 | 99.39 | 92.0 |
