## Appendix 7 for "CohortContrast: An R Package for Enrichment-Based Identification of Clinically Relevant Concepts in OMOP CDM Data"

**Appendix 7: Extracted concepts for lung cancer cohort**

| **Concept name** | **Heritage** | **Target prev.** | **Enrichment** | **MD1** | **MD2** | **LLM** |
| --- | --- | --- | --- | --- | --- | --- |
| Nausea and vomiting | condition_occurrence | 0.022 | 6.752 | −1 | 0 | 0 |
| Dysphagia | condition_occurrence | 0.012 | 3.993 | 0 | −1 | 0 |
| Constipation | condition_occurrence | 0.025 | 2.243 | −1 | −1 | 0 |
| Chest pain | condition_occurrence | 0.023 | 2.367 | +1 | +1 | 0 |
| Urinary tract infectious disease | condition_occurrence | 0.060 | 2.777 | −1 | −1 | 0 |
| Cachexia | condition_occurrence | 0.038 | 65.844 | 0 | +1 | +1 |
| Retention of urine | condition_occurrence | 0.020 | 3.898 | −1 | −1 | 0 |
| Acute kidney injury | condition_occurrence | 0.024 | 12.829 | −1 | −1 | 0 |
| Chronic renal failure | condition_occurrence | 0.037 | 2.804 | −1 | 0 | 0 |
| Cough | condition_occurrence | 0.028 | 2.625 | +1 | 0 | 0 |
| Hemoptysis | condition_occurrence | 0.034 | 19.366 | +1 | +1 | +1 |
| Old myocardial infarction | condition_occurrence | 0.102 | 1.424 | −1 | 0 | −1 |
| Atherosclerosis of arteries of the extremities | condition_occurrence | 0.089 | 1.424 | −1 | 0 | −1 |
| Coronary arteriosclerosis | condition_occurrence | 0.049 | 1.615 | −1 | 0 | −1 |
| Hypertensive heart disease without congestive heart failure | condition_occurrence | 0.390 | 1.414 | −1 | −1 | −1 |
| Hypo-osmolality and or hyponatremia | condition_occurrence | 0.022 | 29.892 | 0 | 0 | +1 |
| Sleep disorder | condition_occurrence | 0.080 | 1.522 | −1 | −1 | 0 |
| Hypokalemia | condition_occurrence | 0.013 | 15.493 | −1 | 0 | 0 |
| Pulmonary embolism | condition_occurrence | 0.054 | 10.148 | 0 | +1 | 0 |
| Agranulocytosis | condition_occurrence | 0.040 | 55.774 | 0 | +1 | 0 |
| Anemia in neoplastic disease | condition_occurrence | 0.118 | 36.950 | +1 | +1 | +1 |
| Complication of procedure | condition_occurrence | 0.012 | 2.704 | 0 | 0 | 0 |
| Cerebral infarction | condition_occurrence | 0.023 | 2.086 | −1 | 0 | 0 |
| Disorder due to type 2 diabetes mellitus | condition_occurrence | 0.092 | 1.787 | −1 | −1 | −1 |
| Atrial arrhythmia | condition_occurrence | 0.206 | 2.197 | −1 | 0 | 0 |
| Sequelae of tuberculosis | condition_occurrence | 0.017 | 2.062 | 0 | 0 | −1 |
| Sequelae of cerebral infarction | condition_occurrence | 0.051 | 1.992 | −1 | 0 | 0 |
| Chronic intractable pain | condition_occurrence | 0.016 | 3.645 | 0 | −1 | 0 |
| Aortic valve stenosis | condition_occurrence | 0.018 | 2.181 | −1 | 0 | −1 |
| Thrombophlebitis of deep veins of lower extremity | condition_occurrence | 0.014 | 2.838 | −1 | +1 | 0 |
| Disorder of vein | condition_occurrence | 0.012 | 2.856 | 0 | 0 | 0 |
| Pressure injury | condition_occurrence | 0.027 | 12.333 | −1 | −1 | 0 |
| COVID-19 | condition_occurrence | 0.032 | 219.631 | −1 | −1 | 0 |
| Secondary thrombocytopenia | condition_occurrence | 0.020 | 23.239 | 0 | +1 | 0 |
| Chronic kidney disease | condition_occurrence | 0.028 | 3.038 | −1 | 0 | 0 |
| Neoplastic disease of uncertain behavior | condition_occurrence | 0.022 | 9.569 | +1 | −1 | 0 |
| Sepsis | condition_occurrence | 0.078 | 15.788 | 0 | 0 | 0 |
| Malignant neoplastic disease | condition_occurrence | 0.046 | 45.567 | +1 | 0 | +1 |
| Heart failure | condition_occurrence | 0.377 | 1.756 | 0 | −1 | 0 |
| Bacterial infectious disease | condition_occurrence | 0.056 | 16.721 | 0 | −1 | 0 |
| Anemia | condition_occurrence | 0.099 | 3.665 | 0 | 0 | 0 |
| Malignant tumor of lung | condition_occurrence | 0.997 | 861.211 | +1 | +1 | +1 |
| Disorder of respiratory system | condition_occurrence | 0.854 | 4.263 | +1 | 0 | +1 |
| Focal onset impaired awareness epileptic seizure + Localization related symptomatic epilepsy | condition_occurrence | 0.015 | 3.322 | +1 | 0 | 0 |
| Death Certificate | death | 0.478 | 100.000 | 0 | 0 | +1 |
| quetiapine 25 MG Oral Tablet | drug_exposure | 0.040 | 3.768 | −1 | −1 | 0 |
| Fenoterol 0.05 MG/ACTUAT / Ipratropium 0.02 MG/ACTUAT Inhalation Solution | drug_exposure | 0.053 | 2.932 | 0 | 0 | 0 |
| esomeprazole 20 MG Oral Capsule | drug_exposure | 0.024 | 3.147 | −1 | −1 | 0 |
| metoclopramide 10 MG Oral Tablet | drug_exposure | 0.250 | 36.686 | 0 | +1 | 0 |
| torsemide 10 MG Oral Tablet | drug_exposure | 0.122 | 2.904 | 0 | −1 | 0 |
| torsemide 5 MG Oral Tablet | drug_exposure | 0.019 | 3.146 | 0 | −1 | 0 |
| furosemide 60 MG Extended Release Oral Capsule | drug_exposure | 0.027 | 1.814 | 0 | −1 | 0 |
| furosemide 40 MG Oral Tablet | drug_exposure | 0.019 | 2.271 | 0 | −1 | 0 |
| tramadol hydrochloride 50 MG Oral Capsule | drug_exposure | 0.044 | 4.930 | 0 | 0 | 0 |
| ketoprofen 50 MG Oral Tablet | drug_exposure | 0.010 | 5.848 | 0 | 0 | 0 |
| ketoprofen 100 MG Oral Tablet | drug_exposure | 0.036 | 5.765 | 0 | 0 | 0 |
| theophylline 200 MG Extended Release Oral Tablet | drug_exposure | 0.056 | 2.828 | 0 | 0 | 0 |
| filgrastim | drug_exposure | 0.150 | 61.113 | 0 | +1 | +1 |
| albumin human, USP | drug_exposure | 0.061 | 13.582 | 0 | −1 | 0 |
| etoposide 100 MG Oral Capsule | drug_exposure | 0.036 | 100.000 | +1 | +1 | +1 |
| dexamethasone 0.5 MG Oral Tablet | drug_exposure | 0.092 | 70.882 | 0 | +1 | 0 |
| dexamethasone 4 MG Oral Tablet | drug_exposure | 0.135 | 935.029 | 0 | +1 | 0 |
| prednisolone 5 MG Oral Tablet | drug_exposure | 0.127 | 7.100 | 0 | +1 | 0 |
| amoxicillin 500 MG / clavulanate 125 MG Oral Tablet | drug_exposure | 0.023 | 2.781 | 0 | 0 | 0 |
| clarithromycin 500 MG Oral Tablet | drug_exposure | 0.068 | 1.384 | 0 | 0 | 0 |
| albuterol 0.1 MG/ACTUAT Inhalation Suspension | drug_exposure | 0.055 | 2.128 | 0 | 0 | 0 |
| ibuprofen 400 MG Oral Tablet | drug_exposure | 0.018 | 3.374 | 0 | 0 | 0 |
| ibuprofen 600 MG Oral Tablet | drug_exposure | 0.065 | 2.762 | 0 | 0 | 0 |
| torsemide 100 MG Oral Tablet | drug_exposure | 0.020 | 2.409 | 0 | 0 | 0 |
| cefuroxime 500 MG Oral Tablet | drug_exposure | 0.118 | 4.693 | 0 | 0 | 0 |
| ciprofloxacin 500 MG Oral Tablet | drug_exposure | 0.079 | 1.835 | 0 | 0 | 0 |
| gabapentin 300 MG Oral Capsule | drug_exposure | 0.105 | 5.128 | 0 | 0 | 0 |
| granisetron 1 MG Oral Tablet | drug_exposure | 0.195 | 53.951 | 0 | +1 | +1 |
| spironolactone 25 MG Oral Tablet | drug_exposure | 0.031 | 2.102 | 0 | 0 | 0 |
| esomeprazole 40 MG Oral Tablet | drug_exposure | 0.039 | 1.860 | 0 | 0 | 0 |
| esomeprazole 40 MG Oral Capsule | drug_exposure | 0.020 | 2.199 | 0 | 0 | 0 |
| tramadol 100 MG Oral Tablet | drug_exposure | 0.142 | 4.962 | 0 | +1 | 0 |
| pregabalin 75 MG Oral Capsule | drug_exposure | 0.022 | 2.923 | 0 | 0 | 0 |
| miconazole 0.02 MG/MG Oral Gel | drug_exposure | 0.018 | 4.994 | −1 | 0 | 0 |
| amoxicillin 875 MG / clavulanate 125 MG Oral Tablet | drug_exposure | 0.228 | 4.182 | 0 | 0 | 0 |
| tramadol hydrochloride 50 MG Oral Tablet | drug_exposure | 0.014 | 3.544 | 0 | 0 | 0 |
| olodaterol 0.0025 MG/ACTUAT / tiotropium 0.0025 MG/ACTUAT Inhalant Solution | drug_exposure | 0.032 | 6.249 | 0 | 0 | 0 |
| Tranexamic Acid 500 MG/ML Oral Solution | drug_exposure | 0.037 | 19.489 | −1 | 0 | 0 |
| tiotropium 0.0025 MG/ACTUAT Inhalant Solution | drug_exposure | 0.041 | 1.686 | 0 | 0 | 0 |
| fluticasone 0.184 MG/ACTUAT / vilanterol 0.022 MG/ACTUAT Inhalation Powder | drug_exposure | 0.012 | 3.190 | 0 | 0 | 0 |
| glycopyrronium 0.044 MG / indacaterol 0.085 MG Inhalant Powder | drug_exposure | 0.017 | 2.776 | 0 | 0 | 0 |
| umeclidinium 0.055 MG/ACTUAT / vilanterol 0.022 MG/ACTUAT Inhalant Powder | drug_exposure | 0.030 | 5.444 | 0 | 0 | 0 |
| bemiparin 25000 UNT/ML Injectable Solution | drug_exposure | 0.022 | 29.709 | 0 | 0 | 0 |
| metoprolol succinate 25 MG Extended Release Oral Tablet | drug_exposure | 0.059 | 1.456 | 0 | 0 | −1 |
| morphine sulfate 10 MG Oral Tablet | drug_exposure | 0.103 | 354.509 | 0 | +1 | 0 |
| morphine sulfate 20 MG Oral Tablet | drug_exposure | 0.022 | 77.463 | 0 | +1 | 0 |
| morphine sulfate 30 MG Extended Release Oral Capsule | drug_exposure | 0.029 | 202.316 | 0 | +1 | 0 |
| morphine sulfate 30 MG Extended Release Oral Tablet | drug_exposure | 0.022 | 152.193 | 0 | +1 | 0 |
| acetaminophen 500 MG / codeine phosphate 30 MG Oral Tablet | drug_exposure | 0.348 | 8.284 | 0 | 0 | 0 |
| oxycodone hydrochloride 10 MG Extended Release Oral Tablet | drug_exposure | 0.064 | 55.022 | 0 | +1 | 0 |
| oxycodone hydrochloride 10 MG Oral Tablet | drug_exposure | 0.034 | 118.018 | 0 | +1 | 0 |
| oxycodone hydrochloride 20 MG Extended Release Oral Tablet | drug_exposure | 0.039 | 67.439 | 0 | +1 | 0 |
| oxycodone hydrochloride 40 MG Extended Release Oral Tablet | drug_exposure | 0.011 | 77.463 | 0 | +1 | 0 |
| oxycodone hydrochloride 5 MG Oral Tablet | drug_exposure | 0.036 | 123.486 | 0 | +1 | 0 |
| rivaroxaban 15 MG Oral Tablet | drug_exposure | 0.026 | 3.556 | 0 | +1 | 0 |
| rivaroxaban 20 MG Oral Tablet | drug_exposure | 0.044 | 3.289 | 0 | +1 | 0 |
| fluticasone 0.093 MG/ACTUAT / vilanterol 0.022 MG/ACTUAT Inhalant Powder | drug_exposure | 0.013 | 2.654 | 0 | 0 | 0 |
| Enoxaparin 10000 UNT/ML Prefilled Syringe | drug_exposure | 0.026 | 8.243 | 0 | +1 | 0 |
| pantoprazole 20 MG Oral Tablet | drug_exposure | 0.062 | 2.276 | 0 | 0 | 0 |
| pantoprazole 40 MG Oral Tablet | drug_exposure | 0.132 | 3.755 | 0 | 0 | 0 |
| dihydrocodeine bitartrate 60 MG Extended Release Oral Tablet | drug_exposure | 0.014 | 8.782 | 0 | +1 | 0 |
| fentanyl 0.025 MG/HR Transdermal System | drug_exposure | 0.018 | 100.000 | 0 | +1 | 0 |
| fentanyl 0.05 MG/HR Transdermal System | drug_exposure | 0.015 | 100.000 | 0 | +1 | 0 |
| sultamicillin 375 MG Oral Tablet | drug_exposure | 0.043 | 5.322 | 0 | 0 | 0 |
| apixaban 2.5 MG Oral Tablet | drug_exposure | 0.024 | 4.943 | 0 | +1 | 0 |
| apixaban 5 MG Oral Tablet | drug_exposure | 0.031 | 4.779 | 0 | +1 | 0 |
| aclidinium 0.322 MG/ACTUAT Inhalant Powder | drug_exposure | 0.018 | 2.412 | 0 | 0 | 0 |
| aclidinium 0.34 MG/ACTUAT / formoterol 0.012 MG/ACTUAT Inhalant Powder | drug_exposure | 0.015 | 6.436 | 0 | 0 | 0 |
| Omeprazole 20 MG Oral Capsule | drug_exposure | 0.191 | 1.896 | 0 | 0 | 0 |
| Omeprazole 40 MG Oral Capsule | drug_exposure | 0.051 | 2.607 | 0 | 0 | 0 |
| Bronchodilatation test | measurement | 0.228 | 4.295 | 0 | +1 | 0 |
| Measurement of Severe acute respiratory syndrome coronavirus 2 (SARS-CoV-2) | measurement | 0.041 | 93.867 | −1 | 0 | 0 |
| Measurement of Severe acute respiratory syndrome coronavirus 2 (SARS-CoV-2) using Nucleic acid amplification technique in Unspecified specimen | measurement | 0.164 | 100.000 | −1 | 0 | 0 |
| AJCC/UICC pathological T1b Category | measurement | 0.039 | 45.263 | +1 | +1 | +1 |
| AJCC/UICC pathological T1a Category | measurement | 0.023 | 52.857 | +1 | +1 | +1 |
| AJCC/UICC clinical T2b Category | measurement | 0.034 | 100.000 | +1 | +1 | +1 |
| AJCC/UICC pathological N2 Category | measurement | 0.017 | 100.000 | +1 | +1 | +1 |
| AJCC/UICC clinical N3 Category | measurement | 0.117 | 806.531 | +1 | +1 | +1 |
| AJCC/UICC pathological T4 Category | measurement | 0.020 | 136.700 | +1 | +1 | +1 |
| AJCC/UICC Stage 2B | measurement | 0.031 | 53.541 | +1 | +1 | +1 |
| AJCC/UICC Stage 2A | measurement | 0.021 | 11.847 | +1 | +1 | +1 |
| AJCC/UICC clinical N0 Category | measurement | 0.083 | 9.231 | +1 | +1 | +1 |
| AJCC/UICC Stage 3C | measurement | 0.014 | 24.606 | +1 | +1 | +1 |
| AJCC/UICC clinical T1c Category | measurement | 0.015 | 51.946 | +1 | +1 | +1 |
| AJCC/UICC pathological T3 Category | measurement | 0.026 | 13.670 | +1 | +1 | +1 |
| AJCC/UICC Stage 3B | measurement | 0.062 | 142.168 | +1 | +1 | +1 |
| AJCC/UICC clinical TX Category | measurement | 0.022 | 100.000 | +1 | +1 | +1 |
| AJCC/UICC Stage 1B | measurement | 0.038 | 52.311 | +1 | +1 | +1 |
| AJCC/UICC clinical N2 Category | measurement | 0.181 | 1,248.528 | +1 | +1 | +1 |
| AJCC/UICC pathological T2a Category | measurement | 0.044 | 300.740 | +1 | +1 | +1 |
| AJCC/UICC clinical T2a Category | measurement | 0.053 | 183.634 | +1 | +1 | +1 |
| AJCC/UICC clinical NX Category | measurement | 0.032 | 22.328 | +1 | +1 | +1 |
| AJCC/UICC pathological T1c Category | measurement | 0.017 | 120.296 | +1 | +1 | +1 |
| AJCC/UICC clinical T2 Category | measurement | 0.025 | 174.065 | +1 | +1 | +1 |
| AJCC/UICC clinical T1b Category | measurement | 0.023 | 158.572 | +1 | +1 | +1 |
| AJCC/UICC clinical M0 Category | measurement | 0.328 | 23.366 | +1 | +1 | +1 |
| AJCC/UICC clinical T4 Category | measurement | 0.145 | 499.411 | +1 | +1 | +1 |
| AJCC/UICC pathological T2b Category | measurement | 0.013 | 92.956 | +1 | +1 | +1 |
| AJCC/UICC pathological N0 Category | measurement | 0.104 | 47.815 | +1 | +1 | +1 |
| AJCC/UICC pathological N1 Category | measurement | 0.022 | 37.820 | +1 | +1 | +1 |
| AJCC/UICC clinical N1 Category | measurement | 0.047 | 80.425 | +1 | +1 | +1 |
| AJCC/UICC Stage 3A | measurement | 0.080 | 100.000 | +1 | +1 | +1 |
| AJCC/UICC clinical T3 Category | measurement | 0.086 | 65.920 | +1 | +1 | +1 |
| Blood group antibody screen.cells I+II+III [Presence] in Serum or Plasma | measurement | 0.557 | 14.571 | 0 | 0 | −1 |
| Indirect antiglobulin test.polyspecific reagent [Presence] in Serum or Plasma | measurement | 0.094 | 2.248 | 0 | 0 | −1 |
| Blood group antibodies identified in Serum or Plasma | measurement | 0.011 | 12.151 | 0 | 0 | −1 |
| Procalcitonin [Mass/volume] in Serum or Plasma | measurement | 0.422 | 16.087 | 0 | 0 | 0 |
| Peripheral blood smear examination, light microscopy | measurement | 0.314 | 5.595 | 0 | 0 | 0 |
| Screening for occult blood in feces | measurement | 0.094 | 2.185 | −1 | 0 | −1 |
| Cytologic test | measurement | 0.604 | 24.972 | 0 | 0 | +1 |
| Immunohistochemistry procedure | measurement | 0.593 | 44.058 | 0 | 0 | +1 |
| Viral nucleic acid assay | measurement | 0.047 | 29.577 | 0 | 0 | 0 |
| Screening for cancer | measurement | 0.072 | 12.417 | 0 | +1 | +1 |
| Evaluation of acid-base balance | measurement | 0.402 | 9.287 | +1 | 0 | 0 |
| Albumin measurement | measurement | 0.501 | 7.747 | 0 | 0 | 0 |
| Histopathology test | measurement | 0.454 | 11.150 | 0 | +1 | +1 |
| Red blood cell test | measurement | 0.169 | 8.286 | 0 | 0 | 0 |
| Hemoglobin variant test | measurement | 0.058 | 8.128 | 0 | 0 | −1 |
| Bilirubin measurement | measurement | 0.736 | 5.241 | 0 | 0 | 0 |
| Microscopic urinalysis | measurement | 0.276 | 2.798 | 0 | 0 | 0 |
| Spirometry | measurement | 0.351 | 5.304 | 0 | +1 | 0 |
| Body fluid analysis | measurement | 0.059 | 27.279 | 0 | 0 | 0 |
| Glucose measurement | measurement | 0.769 | 1.595 | 0 | 0 | 0 |
| Serum protein electrophoresis | measurement | 0.034 | 2.662 | 0 | 0 | 0 |
| Diffusion capacity of lung | measurement | 0.270 | 16.773 | +1 | +1 | 0 |
| Hemoglobin A1c measurement | measurement | 0.315 | 1.272 | −1 | 0 | −1 |
| Cell count and differential, body fluid | measurement | 0.096 | 31.419 | 0 | 0 | 0 |
| Heparin assay | measurement | 0.036 | 30.871 | 0 | 0 | 0 |
| Erythrocyte sedimentation rate measurement | measurement | 0.201 | 1.660 | 0 | 0 | 0 |
| Molecular genetic test | measurement | 0.028 | 10.936 | +1 | 0 | +1 |
| Osmolality | measurement | 0.019 | 13.214 | 0 | 0 | 0 |
| Soluble transferrin receptor test | measurement | 0.092 | 5.103 | 0 | 0 | 0 |
| Antithrombin III assay | measurement | 0.077 | 7.773 | 0 | 0 | 0 |
| Fibrin-fibrinogen split products assay | measurement | 0.424 | 4.726 | 0 | 0 | 0 |
| Polymerase chain reaction analysis | measurement | 0.208 | 8.544 | 0 | 0 | 0 |
| Ethanol measurement | measurement | 0.047 | 2.813 | −1 | −1 | −1 |
| Measurement of level of substance in blood | measurement | 0.050 | 4.134 | −1 | −1 | 0 |
| HIV 1+2 Ab+HIV1 p24 Ag [Presence] in Serum or Plasma by Immunoassay | measurement | 0.214 | 7.645 | −1 | 0 | −1 |
| Test strip urinalysis | measurement | 0.411 | 2.058 | −1 | −1 | 0 |
| Immunology laboratory test | measurement | 0.935 | 1.894 | 0 | 0 | 0 |
| AJCC/UICC Stage 1A | measurement | 0.082 | 113.005 | +1 | +1 | +1 |
| Enzyme measurement | measurement | 0.851 | 2.255 | 0 | −1 | 0 |
| Coagulation pathway screening | measurement | 0.642 | 4.519 | 0 | 0 | 0 |
| CBC W Auto Differential panel - Blood | measurement | 0.800 | 1.798 | 0 | 0 | 0 |
| Metastasis | measurement | 0.095 | 72.805 | +1 | +1 | +1 |
| Microbiology procedure | measurement | 0.690 | 4.202 | 0 | 0 | 0 |
| C-reactive protein measurement + Electrolytes measurement | measurement | 0.883 | 2.028 | 0 | 0 | 0 |
| AJCC/UICC clinical M1 Category + AJCC/UICC Stage 4 | measurement | 0.307 | 124.638 | +1 | +1 | +1 |
| Malaise and fatigue | observation | 0.012 | 2.962 | +1 | 0 | 0 |
| Died in care home | observation | 0.083 | 100.000 | 0 | 0 | 0 |
| Patient meets COVID-19 laboratory diagnostic criteria | observation | 0.032 | 219.631 | −1 | −1 | 0 |
| Palliative care | observation | 0.024 | 32.626 | 0 | +1 | +1 |
| Surgical follow-up | observation | 0.018 | 2.497 | 0 | +1 | 0 |
| Day hospital care | observation | 0.070 | 9.733 | 0 | +1 | 0 |
| Died in hospital | observation | 0.214 | 100.000 | 0 | +1 | 0 |
| Follow-up visit | observation | 0.909 | 1.170 | 0 | 0 | 0 |
| Admission by nurse | observation | 0.336 | 3.481 | 0 | 0 | 0 |
| Admission by general nurse | observation | 0.615 | 1.093 | 0 | 0 | 0 |
| Emergency department patient visit | observation | 0.023 | 19.594 | 0 | 0 | 0 |
| Speech therapy | observation | 0.013 | 2.881 | 0 | 0 | 0 |
| Planned procedure | observation | 0.363 | 3.260 | 0 | 0 | 0 |
| Home care by visiting nurse | observation | 0.121 | 13.100 | 0 | 0 | 0 |
| Home visit | observation | 0.147 | 5.102 | 0 | 0 | 0 |
| Arterial pressure monitoring, invasive method | observation | 0.206 | 9.439 | −1 | −1 | 0 |
| Telephone consultation | observation | 0.691 | 1.465 | 0 | 0 | 0 |
| Preparation of medical certificate | observation | 0.032 | 1.862 | −1 | 0 | −1 |
| Occupational therapy | observation | 0.016 | 2.531 | −1 | 0 | 0 |
| Intracranial hemorrhage or cerebral infarction w CC | observation | 0.013 | 3.443 | −1 | 0 | 0 |
| Other resp system O.R. procedures w CC | observation | 0.050 | 172.698 | 0 | +1 | 0 |
| Other resp system O.R. procedures w/o CC/MCC | observation | 0.027 | 61.363 | 0 | +1 | 0 |
| Respiratory infections & inflammations w CC | observation | 0.021 | 10.285 | 0 | 0 | 0 |
| Pulmonary edema & respiratory failure | observation | 0.013 | 3.255 | 0 | −1 | 0 |
| Simple pneumonia & pleurisy w CC | observation | 0.049 | 9.442 | 0 | 0 | 0 |
| Radiotherapy (2) | observation | 0.118 | 50.807 | +1 | +1 | +1 |
| Coronary artery graft present | observation | 0.058 | 1.466 | −1 | 0 | −1 |
| Immediate resuscitation level emergency care | observation | 0.039 | 13.579 | 0 | 0 | 0 |
| Very urgent level emergency care | observation | 0.080 | 17.287 | 0 | 0 | 0 |
| Standard level emergency care | observation | 0.100 | 5.036 | 0 | 0 | 0 |
| Urgent level emergency care | observation | 0.257 | 9.232 | 0 | 0 | 0 |
| eConsultation via online application | observation | 0.054 | 5.648 | 0 | 0 | 0 |
| Preparation for intensity modulated radiation therapy | observation | 0.016 | 13.898 | +1 | +1 | +1 |
| Rapid immunoassay | observation | 0.220 | 3.822 | 0 | 0 | 0 |
| Admission by physician | observation | 0.975 | 1.038 | 0 | 0 | 0 |
| Death | observation | 0.474 | 100.000 | 0 | +1 | +1 |
| Anesthesia duration + Recovery room monitoring, anesthesia | observation | 0.357 | 4.447 | 0 | +1 | 0 |
| Biopsy of mediastinum | procedure_occurrence | 0.132 | 304.689 | +1 | +1 | +1 |
| Hematoxylin and eosin stain method | procedure_occurrence | 0.542 | 8.688 | 0 | +1 | +1 |
| Quarantine | procedure_occurrence | 0.015 | 103.892 | 0 | 0 | −1 |
| Central venous cannula insertion | procedure_occurrence | 0.052 | 5.842 | 0 | 0 | 0 |
| Long-term oxygen therapy | procedure_occurrence | 0.025 | 14.505 | 0 | +1 | 0 |
| Emergency examination for triage | procedure_occurrence | 0.288 | 2.823 | 0 | 0 | 0 |
| Endoscopy of trachea | procedure_occurrence | 0.160 | 550.901 | +1 | 0 | +1 |
| Indirect encounter | procedure_occurrence | 0.036 | 3.554 | 0 | 0 | 0 |
| Individual psychotherapy | procedure_occurrence | 0.025 | 3.742 | 0 | 0 | 0 |
| 6-minute walk test | procedure_occurrence | 0.015 | 6.379 | 0 | 0 | 0 |
| Angiography of cerebral arteries | procedure_occurrence | 0.019 | 2.376 | −1 | 0 | 0 |
| Home oxygen therapy | procedure_occurrence | 0.050 | 26.359 | 0 | 0 | 0 |
| Neurology service | procedure_occurrence | 0.052 | 5.931 | 0 | 0 | 0 |
| Plain chest X-ray | procedure_occurrence | 0.769 | 3.011 | +1 | +1 | +1 |
| Needle biopsy of lung | procedure_occurrence | 0.029 | 101.158 | +1 | +1 | +1 |
| Treatment planning for external beam radiation therapy | procedure_occurrence | 0.093 | 214.163 | +1 | +1 | +1 |
| Endoscopic procedure | procedure_occurrence | 0.027 | 2.434 | +1 | +1 | 0 |
| Electroencephalogram | procedure_occurrence | 0.011 | 3.190 | 0 | 0 | 0 |
| Mycobacterial microscopy | procedure_occurrence | 0.320 | 23.059 | 0 | 0 | −1 |
| Electrocardiographic monitoring | procedure_occurrence | 0.845 | 2.353 | 0 | 0 | 0 |
| Specialized medical examination | procedure_occurrence | 0.084 | 5.068 | 0 | −1 | 0 |
| Insertion of drain using ultrasound guidance | procedure_occurrence | 0.052 | 11.938 | 0 | +1 | 0 |
| Endoscopic biopsy | procedure_occurrence | 0.420 | 11.342 | +1 | 0 | +1 |
| Echocardiography | procedure_occurrence | 0.242 | 3.174 | 0 | 0 | 0 |
| Physical therapy procedure | procedure_occurrence | 0.278 | 3.850 | 0 | 0 | 0 |
| Thoracentesis | procedure_occurrence | 0.151 | 129.979 | +1 | +1 | 0 |
| Chemotherapy | procedure_occurrence | 0.455 | 32.751 | +1 | +1 | +1 |
| Immunoblot assay | procedure_occurrence | 0.034 | 2.261 | 0 | 0 | 0 |
| US scan of abdomen and pelvis | procedure_occurrence | 0.235 | 1.830 | −1 | 0 | 0 |
| Application of dressing | procedure_occurrence | 0.020 | 3.395 | 0 | 0 | 0 |
| Core needle biopsy | procedure_occurrence | 0.072 | 41.542 | +1 | +1 | +1 |
| Fine needle aspiration biopsy of soft tissue | procedure_occurrence | 0.013 | 9.923 | +1 | +1 | 0 |
| Core needle biopsy using ultrasound guidance | procedure_occurrence | 0.179 | 16.749 | +1 | +1 | +1 |
| Fine needle aspiration biopsy using imaging guidance | procedure_occurrence | 0.067 | 5.222 | +1 | +1 | +1 |
| Speech assessment | procedure_occurrence | 0.020 | 3.717 | 0 | 0 | 0 |
| Attention to sutures | procedure_occurrence | 0.046 | 2.141 | 0 | 0 | 0 |
| Endoscopic ultrasonography guided fine needle aspiration | procedure_occurrence | 0.158 | 181.811 | +1 | +1 | +1 |
| Catheterization | procedure_occurrence | 0.135 | 5.938 | 0 | 0 | 0 |
| Magnetic resonance imaging | procedure_occurrence | 0.111 | 2.571 | +1 | +1 | +1 |
| Ultrasonography | procedure_occurrence | 0.045 | 8.374 | 0 | 0 | 0 |
| Medical examination for suspected condition | procedure_occurrence | 0.226 | 2.606 | 0 | 0 | 0 |
| Plain radiography | procedure_occurrence | 0.166 | 6.808 | 0 | 0 | 0 |
| Drainage of pleural cavity | procedure_occurrence | 0.293 | 134.817 | +1 | +1 | 0 |
| Nuclear medicine procedure | procedure_occurrence | 0.194 | 51.455 | +1 | +1 | 0 |
| Emergency procedure | procedure_occurrence | 0.235 | 3.693 | 0 | 0 | 0 |
| Ultrasonography of soft tissue | procedure_occurrence | 0.234 | 4.554 | 0 | 0 | 0 |
| Counseling | procedure_occurrence | 0.114 | 2.599 | 0 | 0 | 0 |
| Puncture procedure | procedure_occurrence | 0.061 | 1.823 | 0 | +1 | 0 |
| Evaluation of biopsy specimen | procedure_occurrence | 0.719 | 8.417 | +1 | +1 | +1 |
| Bronchoscopy | procedure_occurrence | 0.578 | 49.873 | +1 | +1 | +1 |
| Intubation | procedure_occurrence | 0.060 | 83.114 | 0 | 0 | 0 |
| Administration of medication | procedure_occurrence | 0.139 | 35.508 | 0 | 0 | 0 |
| Endoscopic operation | procedure_occurrence | 0.092 | 3.103 | +1 | 0 | 0 |
| Insertion of tracheostomy tube + Surgical procedure | procedure_occurrence | 0.378 | 21.049 | 0 | 0 | 0 |
| Insertion of catheter into urinary bladder + Insertion of indwelling catheter into urinary bladder | procedure_occurrence | 0.252 | 5.950 | 0 | 0 | 0 |
| Endoscopy of stomach + Esophagogastroduodenoscopy | procedure_occurrence | 0.131 | 2.233 | −1 | 0 | −1 |
| Developing a treatment plan + Multidisciplinary cancer case management | procedure_occurrence | 0.704 | 55.885 | +1 | +1 | +1 |
| Radiotherapy + Three dimensional treatment planning for external beam radiation therapy | procedure_occurrence | 0.253 | 38.762 | +1 | +1 | +1 |
| Analysis using real time PCR + Chromosome analysis, cytogenetic procedure AND/OR molecular biology method | procedure_occurrence | 0.382 | 11.786 | +1 | +1 | +1 |
| Computed tomography + Radiology of two body areas | procedure_occurrence | 0.918 | 5.419 | +1 | +1 | +1 |
| Outpatient Visit | visit_detail | 0.985 | 1.016 | 0 | 0 | 0 |
| Pharmacy visit | visit_detail | 0.938 | 1.112 | 0 | 0 | 0 |
| Inpatient Visit | visit_detail | 0.899 | 5.272 | 0 | 0 | 0 |
| Inpatient Visit (visit_occurrence) | visit_occurrence | 0.877 | 5.296 | 0 | 0 | 0 |
| Outpatient Visit (visit_occurrence) | visit_occurrence | 0.980 | 1.022 | 0 | 0 | 0 |
| Pharmacy visit (visit_occurrence) | visit_occurrence | 0.938 | 1.112 | 0 | 0 | 0 |
