## Appendix 8 for "CohortContrast: An R Package for Enrichment-Based Identification of Clinically Relevant Concepts in OMOP CDM Data"

**Appendix 8: Extracted concepts for the prostate cancer cohort**

| **Concept name** | **Heritage** | **Target prev.** | **Enrichment** | **MD1** | **MD2** | **LLM** |
| --- | --- | --- | --- | --- | --- | --- |
| Hyperplasia of prostate | condition_occurrence | 0.566 | 1.940 | 0 | 0 | 0 |
| Inflammatory disorder of male genital organ | condition_occurrence | 0.010 | 4.118 | 0 | 0 | −1 |
| Primary malignant neoplasm of prostate | condition_occurrence | 0.999 | 280.328 | +1 | +1 | +1 |
| Hypertensive heart disease without congestive heart failure | condition_occurrence | 0.353 | 1.336 | −1 | −1 | 0 |
| Essential hypertension | condition_occurrence | 0.330 | 1.221 | −1 | −1 | 0 |
| Complication of medical care | condition_occurrence | 0.018 | 38.582 | 0 | 0 | 0 |
| Complication of procedure | condition_occurrence | 0.013 | 6.125 | 0 | 0 | 0 |
| Adenocarcinoma of prostate | condition_occurrence | 0.481 | 2,565.507 | +1 | +1 | +1 |
| Inguinal hernia | condition_occurrence | 0.025 | 1.691 | −1 | −1 | −1 |
| COVID-19 | condition_occurrence | 0.034 | 181.833 | −1 | −1 | −1 |
| Secondary erectile dysfunction | condition_occurrence | 0.017 | 4.712 | 0 | +1 | 0 |
| Urinary incontinence | condition_occurrence | 0.025 | 12.746 | 0 | +1 | 0 |
| Disorder of prostate | condition_occurrence | 0.370 | 4.109 | +1 | 0 | +1 |
| Disorder of the urinary system | condition_occurrence | 0.218 | 5.396 | +1 | 0 | 0 |
| SARS-COV-2 (COVID-19) vaccine, UNSPECIFIED | drug_exposure | 0.131 | 100.000 | −1 | −1 | −1 |
| nitrofurantoin 50 MG Oral Tablet | drug_exposure | 0.032 | 4.269 | 0 | 0 | 0 |
| triptorelin | drug_exposure | 0.012 | 100.000 | +1 | +1 | +1 |
| albumin human, USP | drug_exposure | 0.055 | 24.537 | 0 | 0 | 0 |
| bicalutamide 50 MG Oral Tablet | drug_exposure | 0.034 | 100.000 | +1 | +1 | +1 |
| trimethoprim 100 MG Oral Tablet | drug_exposure | 0.013 | 9.626 | 0 | 0 | 0 |
| metronidazole 500 MG Oral Tablet | drug_exposure | 0.029 | 1.618 | 0 | 0 | 0 |
| iodine | drug_exposure | 0.139 | 212.029 | −1 | 0 | 0 |
| ciprofloxacin 250 MG Oral Tablet | drug_exposure | 0.014 | 3.181 | 0 | 0 | 0 |
| sulfamethoxazole 800 MG / trimethoprim 160 MG Oral Tablet | drug_exposure | 0.039 | 3.263 | 0 | 0 | 0 |
| norfloxacin 400 MG Oral Tablet | drug_exposure | 0.025 | 2.968 | 0 | 0 | 0 |
| ofloxacin 200 MG Oral Tablet | drug_exposure | 0.036 | 9.645 | 0 | 0 | 0 |
| cefuroxime 500 MG Oral Tablet | drug_exposure | 0.107 | 4.811 | 0 | 0 | 0 |
| ciprofloxacin 500 MG Oral Tablet | drug_exposure | 0.454 | 8.746 | 0 | 0 | 0 |
| goserelin 3.6 MG Drug Implant | drug_exposure | 0.031 | 100.000 | +1 | +1 | +1 |
| bicalutamide 150 MG Oral Tablet | drug_exposure | 0.266 | 1,415.898 | +1 | +1 | +1 |
| Amlodipine 5 MG / Indapamide 1.25 MG / Perindopril 5 MG Oral Tablet | drug_exposure | 0.014 | 2.267 | −1 | 0 | 0 |
| diclofenac sodium 100 MG Rectal Suppository | drug_exposure | 0.010 | 2.400 | 0 | +1 | 0 |
| tolterodine tartrate 2 MG Oral Tablet | drug_exposure | 0.010 | 9.985 | 0 | +1 | 0 |
| tolterodine tartrate 4 MG Extended Release Oral Capsule | drug_exposure | 0.029 | 20.860 | 0 | +1 | 0 |
| tamsulosin hydrochloride 0.4 MG Oral Capsule | drug_exposure | 0.219 | 1.944 | +1 | 0 | 0 |
| rivaroxaban 10 MG Oral Tablet | drug_exposure | 0.014 | 2.512 | −1 | 0 | 0 |
| Enoxaparin 10000 UNT/ML Prefilled Syringe | drug_exposure | 0.098 | 47.577 | −1 | +1 | 0 |
| mirabegron 50 MG Extended Release Oral Tablet | drug_exposure | 0.021 | 10.195 | 0 | 0 | 0 |
| sultamicillin 375 MG Oral Tablet | drug_exposure | 0.041 | 4.694 | 0 | 0 | 0 |
| tamsulosin 0.4 MG Oral Tablet | drug_exposure | 0.049 | 2.334 | +1 | 0 | 0 |
| Fluocortolone 1 MG / Lidocaine 40 MG Rectal Suppository | drug_exposure | 0.028 | 1.918 | −1 | −1 | −1 |
| Fluocortolone 0.001 MG/MG / Lidocaine 0.02 MG/MG Rectal Cream | drug_exposure | 0.014 | 2.117 | −1 | −1 | −1 |
| Triptorelin 3.75 MG Injectable Suspension | drug_exposure | 0.013 | 100.000 | +1 | +1 | +1 |
| Triptorelin 11.25 MG Injectable Suspension | drug_exposure | 0.140 | 100.000 | +1 | +1 | +1 |
| Measurement of Severe acute respiratory syndrome coronavirus 2 (SARS-CoV-2) | measurement | 0.022 | 76.610 | −1 | 0 | −1 |
| Measurement of Severe acute respiratory syndrome coronavirus 2 (SARS-CoV-2) using Nucleic acid amplification technique in Unspecified specimen | measurement | 0.095 | 100.000 | −1 | 0 | −1 |
| AJCC/UICC pathological T3a Category | measurement | 0.046 | 491.964 | +1 | +1 | +1 |
| AJCC/UICC Stage 1 | measurement | 0.167 | 63.589 | +1 | +1 | +1 |
| AJCC/UICC pathological T3b Category | measurement | 0.025 | 100.000 | +1 | +1 | +1 |
| AJCC/UICC Stage 3 | measurement | 0.107 | 1,141.764 | +1 | +1 | +1 |
| AJCC/UICC clinical T2b Category | measurement | 0.024 | 100.000 | +1 | +1 | +1 |
| AJCC/UICC clinical N0 Category | measurement | 0.345 | 76.533 | +1 | +1 | +1 |
| AJCC/UICC Stage 2 | measurement | 0.205 | 242.957 | +1 | +1 | +1 |
| AJCC/UICC clinical T1c Category | measurement | 0.069 | 734.716 | +1 | +1 | +1 |
| AJCC/UICC pathological T2c Category | measurement | 0.093 | 987.621 | +1 | +1 | +1 |
| AJCC/UICC clinical T2c Category | measurement | 0.063 | 100.000 | +1 | +1 | +1 |
| AJCC/UICC clinical T2a Category | measurement | 0.031 | 167.526 | +1 | +1 | +1 |
| AJCC/UICC clinical NX Category | measurement | 0.124 | 120.159 | +1 | +1 | +1 |
| AJCC/UICC clinical T2 Category | measurement | 0.020 | 107.992 | +1 | +1 | +1 |
| AJCC/UICC clinical M0 Category | measurement | 0.521 | 73.076 | +1 | +1 | +1 |
| AJCC/UICC clinical T4 Category | measurement | 0.021 | 100.000 | +1 | +1 | +1 |
| AJCC/UICC pathological N0 Category | measurement | 0.068 | 80.404 | +1 | +1 | +1 |
| AJCC/UICC pathological N1 Category | measurement | 0.012 | 42.766 | +1 | +1 | +1 |
| AJCC/UICC clinical N1 Category | measurement | 0.049 | 104.485 | +1 | +1 | +1 |
| Blood group antibody screen.cells I+II+III [Presence] in Serum or Plasma | measurement | 0.342 | 12.609 | 0 | 0 | 0 |
| Indirect antiglobulin test.polyspecific reagent [Presence] in Serum or Plasma | measurement | 0.141 | 4.644 | 0 | 0 | 0 |
| Procalcitonin [Mass/volume] in Serum or Plasma | measurement | 0.115 | 6.500 | 0 | 0 | 0 |
| Peripheral blood smear examination, light microscopy | measurement | 0.114 | 3.424 | 0 | 0 | 0 |
| Antimicrobial susceptibility test | measurement | 0.051 | 6.675 | 0 | 0 | 0 |
| Aerobic microbial culture | measurement | 0.506 | 5.349 | 0 | 0 | 0 |
| Immunohistochemistry procedure | measurement | 0.406 | 33.003 | 0 | 0 | +1 |
| Evaluation of acid-base balance | measurement | 0.133 | 5.988 | 0 | 0 | 0 |
| Albumin measurement | measurement | 0.336 | 6.523 | 0 | 0 | 0 |
| Histopathology test | measurement | 0.214 | 7.340 | 0 | 0 | +1 |
| Bilirubin measurement | measurement | 0.426 | 3.793 | 0 | 0 | 0 |
| Microscopic urinalysis | measurement | 0.253 | 3.167 | 0 | 0 | 0 |
| Glucose measurement | measurement | 0.725 | 1.514 | 0 | 0 | 0 |
| Hemoglobin A1c measurement | measurement | 0.348 | 1.433 | −1 | 0 | 0 |
| Microbial culture | measurement | 0.098 | 4.481 | 0 | 0 | 0 |
| Uroflowmetry | measurement | 0.047 | 1.846 | +1 | 0 | 0 |
| Fibrin-fibrinogen split products assay | measurement | 0.192 | 3.093 | 0 | 0 | 0 |
| HIV 1+2 Ab+HIV1 p24 Ag [Presence] in Serum or Plasma by Immunoassay | measurement | 0.083 | 3.285 | −1 | 0 | −1 |
| Screening for malignant neoplasm of intestinal tract | measurement | 0.055 | 1.666 | −1 | 0 | −1 |
| Test strip urinalysis | measurement | 0.445 | 2.380 | 0 | 0 | 0 |
| Immunology laboratory test | measurement | 0.928 | 1.814 | 0 | 0 | 0 |
| AJCC/UICC clinical T1 Category | measurement | 0.028 | 27.019 | +1 | +1 | +1 |
| AJCC/UICC clinical T3 Category | measurement | 0.092 | 195.678 | +1 | +1 | +1 |
| Enzyme measurement | measurement | 0.674 | 1.968 | 0 | 0 | 0 |
| Screening for cancer | measurement | 0.043 | 5.144 | 0 | +1 | +1 |
| Coagulation pathway screening | measurement | 0.371 | 3.145 | 0 | 0 | 0 |
| CBC W Auto Differential panel - Blood | measurement | 0.747 | 1.782 | 0 | 0 | 0 |
| AJCC/UICC pathological T2 Category | measurement | 0.058 | 41.228 | +1 | +1 | +1 |
| AJCC/UICC pathological T1 Category | measurement | 0.024 | 11.454 | +1 | +1 | +1 |
| Disk diffusion susceptibility test + Microbial identification test | measurement | 0.187 | 4.965 | 0 | 0 | 0 |
| C-reactive protein measurement + Electrolytes measurement | measurement | 0.762 | 1.973 | 0 | 0 | 0 |
| AJCC/UICC clinical M1 Category + AJCC/UICC Stage 4 | measurement | 0.101 | 179.679 | +1 | +1 | +1 |
| Neurogenic urinary bladder | observation | 0.039 | 6.789 | 0 | 0 | 0 |
| Blood in urine | observation | 0.028 | 6.860 | +1 | +1 | 0 |
| Patient meets COVID-19 laboratory diagnostic criteria | observation | 0.034 | 181.833 | −1 | −1 | −1 |
| Low dose rate brachytherapy | observation | 0.014 | 100.000 | +1 | +1 | +1 |
| Surgical follow-up | observation | 0.018 | 2.743 | 0 | +1 | 0 |
| Day hospital care | observation | 0.038 | 6.234 | 0 | 0 | 0 |
| Follow-up visit | observation | 0.973 | 1.294 | 0 | 0 | 0 |
| Admission by nurse | observation | 0.372 | 3.517 | 0 | 0 | 0 |
| Admission by general nurse | observation | 0.753 | 1.307 | 0 | 0 | 0 |
| Planned procedure | observation | 0.448 | 4.286 | 0 | 0 | 0 |
| Arterial pressure monitoring, invasive method | observation | 0.218 | 17.727 | −1 | 0 | 0 |
| Telephone consultation | observation | 0.706 | 1.528 | 0 | 0 | 0 |
| Preparation of medical certificate | observation | 0.028 | 1.863 | −1 | −1 | 0 |
| Follow-up encounter | observation | 0.092 | 5.351 | −1 | 0 | 0 |
| Requires vaccination | observation | 0.135 | 59.880 | −1 | −1 | −1 |
| Major male pelvic procedures w CC/MCC | observation | 0.066 | 100.000 | +1 | +1 | +1 |
| Major male pelvic procedures w/o CC/MCC | observation | 0.225 | 100.000 | +1 | +1 | +1 |
| Transurethral prostatectomy w CC/MCC | observation | 0.019 | 40.243 | +1 | +1 | 0 |
| Transurethral prostatectomy w/o CC/MCC | observation | 0.026 | 39.953 | +1 | +1 | 0 |
| Malignancy, male reproductive system w CC | observation | 0.050 | 100.000 | +1 | +1 | +1 |
| Malignancy, male reproductive system w/o CC/MCC | observation | 0.060 | 100.000 | +1 | +1 | +1 |
| Radiotherapy (2) | observation | 0.086 | 305.516 | +1 | +1 | +1 |
| Urinary bladder stoma present | observation | 0.044 | 21.397 | +1 | +1 | 0 |
| Standard level emergency care | observation | 0.085 | 3.594 | 0 | 0 | 0 |
| Urgent level emergency care | observation | 0.114 | 5.348 | 0 | 0 | 0 |
| eConsultation via online application | observation | 0.056 | 4.673 | 0 | 0 | 0 |
| Preparation for intensity modulated radiation therapy | observation | 0.150 | 799.788 | +1 | +1 | +1 |
| Admission by physician | observation | 0.996 | 1.061 | 0 | 0 | 0 |
| Anesthesia duration + Recovery room monitoring, anesthesia | observation | 0.489 | 6.989 | 0 | 0 | 0 |
| Chromosome analysis, cytogenetic procedure AND/OR molecular biology method | procedure_occurrence | 0.106 | 4.382 | +1 | 0 | 0 |
| Three dimensional treatment planning for external beam radiation therapy | procedure_occurrence | 0.067 | 178.833 | +1 | +1 | +1 |
| Central venous cannula insertion | procedure_occurrence | 0.024 | 4.772 | 0 | 0 | 0 |
| Plain X-ray abdomen | procedure_occurrence | 0.022 | 2.073 | −1 | −1 | 0 |
| Preventive procedure | procedure_occurrence | 0.015 | 18.050 | −1 | −1 | 0 |
| Medical counseling | procedure_occurrence | 0.062 | 3.081 | −1 | 0 | 0 |
| Emergency procedure | procedure_occurrence | 0.164 | 2.841 | 0 | −1 | 0 |
| Plain chest X-ray | procedure_occurrence | 0.365 | 1.966 | −1 | −1 | 0 |
| Electrocardiographic monitoring | procedure_occurrence | 0.672 | 2.082 | −1 | −1 | 0 |
| Change of cystostomy tube | procedure_occurrence | 0.047 | 16.583 | +1 | 0 | 0 |
| Specialized medical examination | procedure_occurrence | 0.081 | 8.048 | 0 | 0 | 0 |
| Fluoroscopy | procedure_occurrence | 0.019 | 13.414 | 0 | +1 | 0 |
| Intubation | procedure_occurrence | 0.019 | 25.960 | 0 | 0 | 0 |
| Counseling | procedure_occurrence | 0.058 | 1.924 | 0 | 0 | 0 |
| Transurethral cystoscopy | procedure_occurrence | 0.068 | 5.683 | +1 | +1 | 0 |
| US scan of abdomen and pelvis | procedure_occurrence | 0.244 | 1.842 | 0 | +1 | 0 |
| Endoscopy | procedure_occurrence | 0.029 | 23.998 | 0 | 0 | 0 |
| Core needle biopsy | procedure_occurrence | 0.021 | 10.461 | +1 | +1 | +1 |
| Endorectal ultrasonography | procedure_occurrence | 0.455 | 5.053 | +1 | +1 | +1 |
| Fluoroscopy using contrast | procedure_occurrence | 0.240 | 91.543 | +1 | +1 | 0 |
| Paracentesis of urinary bladder | procedure_occurrence | 0.050 | 18.524 | +1 | +1 | 0 |
| Fine needle aspiration biopsy using imaging guidance | procedure_occurrence | 0.030 | 4.316 | +1 | 0 | +1 |
| Attention to sutures | procedure_occurrence | 0.048 | 2.333 | 0 | 0 | 0 |
| Ultrasonography of urinary bladder for post-void residual volume | procedure_occurrence | 0.229 | 2.717 | +1 | +1 | 0 |
| Radiotherapy | procedure_occurrence | 0.229 | 174.251 | +1 | +1 | +1 |
| Administration of medication | procedure_occurrence | 0.042 | 36.920 | 0 | −1 | 0 |
| Magnetic resonance imaging | procedure_occurrence | 0.159 | 3.468 | +1 | +1 | +1 |
| Medical examination for suspected condition | procedure_occurrence | 0.176 | 2.261 | 0 | −1 | 0 |
| Endoscopic procedure | procedure_occurrence | 0.037 | 3.816 | 0 | +1 | 0 |
| Physical therapy procedure | procedure_occurrence | 0.248 | 3.528 | 0 | −1 | 0 |
| Ultrasonography | procedure_occurrence | 0.194 | 3.001 | 0 | −1 | 0 |
| Nuclear medicine procedure | procedure_occurrence | 0.237 | 64.824 | +1 | +1 | +1 |
| Catheterization + Neurology service | procedure_occurrence | 0.132 | 12.039 | 0 | −1 | 0 |
| Computed tomography + Radiology of two body areas | procedure_occurrence | 0.500 | 4.433 | +1 | −1 | 0 |
| Developing a treatment plan + Multidisciplinary cancer case management | procedure_occurrence | 0.383 | 75.687 | +1 | +1 | +1 |
| Evaluation of biopsy specimen + Hematoxylin and eosin stain method | procedure_occurrence | 0.812 | 10.801 | +1 | +1 | +1 |
| Biopsy of prostate + Core needle biopsy using ultrasound guidance | procedure_occurrence | 0.644 | 26.514 | +1 | +1 | +1 |
| Administration into urinary bladder via intravesical route + Irrigation of urinary bladder | procedure_occurrence | 0.012 | 16.268 | +1 | +1 | 0 |
| Insertion of catheter into urinary bladder + Insertion of indwelling catheter into urinary bladder + Surgical procedure | procedure_occurrence | 0.509 | 11.250 | +1 | +1 | 0 |
| Outpatient Visit | visit_detail | 0.998 | 1.031 | 0 | −1 | 0 |
| Pharmacy visit | visit_detail | 0.967 | 1.151 | 0 | −1 | 0 |
| Inpatient Visit | visit_detail | 0.635 | 4.991 | 0 | −1 | 0 |
| Inpatient Visit (visit_occurrence) | visit_occurrence | 0.630 | 5.069 | 0 | −1 | 0 |
| Outpatient Visit (visit_occurrence) | visit_occurrence | 0.993 | 1.034 | 0 | −1 | 0 |
| Pharmacy visit (visit_occurrence) | visit_occurrence | 0.967 | 1.151 | 0 | −1 | 0 |
