## Appendix 9 for "CohortContrast: An R Package for Enrichment-Based Identification of Clinically Relevant Concepts in OMOP CDM Data"

**Appendix 9: Validation of LLM and MD labeling on the extracted concepts**

| Subset | Rater pair | N | Agreement (%) | κ | 95% CI |
| --- | --- | --- | --- | --- | --- |
| Lung | MD1–MD2 | 296 | 68.6 | 0.445 | 0.355–0.499 |
| Lung | MD1–LLM | 296 | 79.1 | 0.590 | 0.502–0.617 |
| Lung | MD2–LLM | 296 | 67.2 | 0.387 | 0.296–0.449 |
| Prostate | MD1–MD2 | 170 | 74.7 | 0.591 | 0.487–0.618 |
| Prostate | MD1–LLM | 170 | 78.8 | 0.638 | 0.537–0.658 |
| Prostate | MD2–LLM | 170 | 71.2 | 0.513 | 0.405–0.566 |
| Overall | MD1–MD2 | 466 | 70.8 | 0.505 | 0.437–0.571 |
| Overall | MD1–LLM | 466 | 79.0 | 0.612 | 0.546–0.657 |
| Overall | MD2–LLM | 466 | 68.7 | 0.438 | 0.368–0.517 |

**Note: Agreement is percent exact agreement across the three labels (direct/indirect/noise). κ is unweighted Cohen’s kappa; CI is 95%.**
